# Longitudinal patterns of smoking behaviours in adolescence and early adulthood and their association with modifiable and sociodemographic risk factors

**DOI:** 10.1101/2024.06.05.24308474

**Authors:** Alexandria Andrayas, Hannah Jones, Jasmine Khouja, Lindsey Hines, Marcus Munafò, Jon Heron, Hannah Sallis

## Abstract

**Introduction:** Smoking contributes a huge burden on public health; thus, identifying risk factors for smoking remains an important area of research. This study adds to the wealth of existing literature by utilising repeated smoking measures collected in a UK sample of young adults to (a) examine differences between longitudinal smoking behaviours, (b) investigate their association with many risk factors, and c) consider how these associations may change over time.

**Methods:** This study uses longitudinal latent class analysis and 12 repeated measures to derive patterns of smoking in the Avon Longitudinal Study of Parents and Children. The association of these patterns with 402 risk factor measures collected from 0-28 years is then investigated. The selected risk factors include familial and peer factors, lifestyle and sociodemographic factors, mental health, parenthood, adverse childhood experiences and trauma.

**Results:** Five different latent classes of smoking were derived and referred to as non-smoking, short-term smoking, occasional smoking, early-onset smoking, and late-onset smoking. These showed differences in age of onset, frequency, and cessation. Other substance use, and parental and peer substance use, showed the strongest association with smoking patterns. More risk factors were associated with early-onset than late-onset smoking. Many risk factors of regular smoking did not show the same associations with occasional smoking. Fewer measures differentiated late-onset from short-term smoking. Some associations varied depending on the time of measurement or smoking pattern in question.

**Conclusions:** Findings from this study may be used to identify groups of people most vulnerable to more harmful smoking patterns despite being exposed to strong tobacco prevention efforts. This could also help better tailor smoking interventions and improve tobacco control policies.

## Introduction

Following primary studies in the UK^1,2^ and the US Surgeon General’s report^3^, an overwhelming amount of evidence has led to the classification of smoking as the leading preventable cause of morbidity and mortality worldwide^4–6^. Changes to tobacco policy, and increases in smoking intervention and cessation campaigns, have been introduced to improve national and global public health. However smoking remains relatively common among young adults^7^. Most people who smoke began using tobacco in their teens^8^. The average age at smoking cessation tends to occur in mid-life but there is much greater variation in when someone stops smoking versus when they start smoking^9^. Though the younger someone is at the time they attempt to stop, the more likely they are to succeed^10^.

Many risk factors of smoking have been described. Predictors of smoking onset identified in systematic reviews include increased age or school grade, lower socioeconomic position (SEP), poor academic performance, sensation seeking or rebelliousness, intention to smoke in the future, receptivity to tobacco promotion efforts, susceptibility to smoking, and family and peer smoking^11,12^. Similarly, predictors of smoking cessation in youth include not having friends who smoke, resisting peer pressure to smoke, not having intentions to smoke in the future, being older at first use of cigarettes, and having negative beliefs about smoking^13,14^. The role of age of onset is particularly important given that in recent years the age of smoking initiation is shifting into early adulthood^15^. Higher smoking prevalence is also linked with almost every indicator of deprivation or marginalisation, and this contributes massively to health inequalities^16^. While modifiable risk factors present potential interventions, sociodemographic factors can be used to better tailor public health policy and smoking prevention programmes^17^.

Although there is a wealth of literature on the predictors of smoking initiation and cessation, existing research has paid little attention to how associations may vary across different smoking patterns or across the life course. Looking at the age that certain risk factors associate with smoking, and how risk factors differ depending on the smoking pattern in question, can improve these efforts. This is particularly important given the “hardening hypothesis” where, as smoking prevalence declines, the remaining smoking population are increasingly resistant to established interventions^18^. Improvements in cessation campaigns are also required given they have made only a modest reduction to health inequalities linked to smoking^19,20^, and the “tobacco endgame” will need to address the higher smoking rates in marginalised communities^21^. With so many risk factors identified for smoking, improving knowledge of which risk factors are related to more harmful patterns of smoking, characterised by earlier age of onset and more frequent smoking, would help reduce preventable non-communicable diseases and cancers linked to smoking^22^.

The present study uses repeated measures of smoking and risk factors, collected throughout childhood, adolescence, and adulthood, within the Avon Longitudinal Study of Parents and Children (ALSPAC). Longitudinal data can reduce recall bias and are more reliable than data collected retrospectively or at a single timepoint. The latter only gives a snapshot of a person’s smoking behaviour and not how it changes over time. ALSPAC is a valuable resource for understanding why some young adults smoke and others do not given early adulthood is a critical period where smoking behaviours often stabilise or end^23^. By utilising a phenotypically rich birth cohort, the present exploratory study seeks to offer insights through hypothesis generation. Many risk factors have been identified from the literature and this study aims to explore their relationship with longitudinal patterns of smoking during adolescence to young adulthood. The most important risk factors for differing smoking patterns are elucidated, and the age at which these risk factors are most influential is investigated.

## Methods

### Participants

Pregnant women resident in Avon, UK with expected dates of delivery between 1st April 1991 and 31^st^ December 1992 were invited to take part in the study, The initial number of pregnancies enrolled was 14,541 and 13,988 children were alive at 1 year of age. An attempt was made to bolster the initial sample with eligible cases who had failed to join the study originally. The total sample size for analyses using any data collected after the age of seven is 15,447 pregnancies, where 14,901 children were alive at 1 year of age. 14,203 unique mothers were initially enrolled, and 14,833 unique women (Generation 0 or G0 mothers) were enrolled in ALSPAC as of September 2021. Detailed information has been collected on these women and their offspring at regular intervals^24–26^. 12,113 G0 partners have been in contact with the study by providing data or formally enrolling when this started in 2010. 3,807 G0 partners are currently enrolled^27,28^. Study data were collected and managed using REDCap electronic data capture tools hosted at the University of Bristol^29^. The study website contains details of all available data: http://www.bristol.ac.uk/alspac/researchers/our-data/.

### Smoking measures

The measures of smoking status and frequency used in this study (Supplementary Text 1) were collected in ALSPAC regularly from 13 to 28 years. Smoking at each timepoint was parameterised as a 3-category variable where each participant was classified by either non-smoking, occasional smoking, or regular smoking.

This analysis was restricted to participants who provided data on smoking in at least three timepoints, including at least one measure in adolescence (13, 14, 15, 16 years; y), young adulthood (17, 18, 20, 21y), and in their twenties (22, 23, 24, 28y). In total 4,937 participants met this criterion. More information on the smoking measures used in this study can be found in Supplementary Tables S1-S2.

### Risk factor measures

A search for ALSPAC measures related to any risk factors of interest (Supplementary Text 2) was carried out using the ALSPAC online variable search tool, the variable catalogue, and the data dictionary. This search included measures from questionnaires (mother- and participant-reported), clinics, biological samples, and other derived variables. A full list of investigated risk factor measures, including when each was collected, and how they were parameterised can be found in Supplementary Tables S6-S7 and S15. All variables were recoded as binary measures if not already.

In total 525 variables related to approximately 95 different risk factors were identified spanning from 0 to 28 years. Supplementary Tables S12-S13 shows the number of repeated measures available per risk factor. Due to sample size constraints (Supplementary Figure S1) 123 measures were not analysed leaving 402 to be investigated. Total sample sizes for the included measures ranged from 796 to 4,930. The number of exposed or unexposed participants for each included measure ranged from 112 to 4,290.

### Data cleaning and software

Data were cleaned and processed in Stata (version 17)^30^ removing any multiple births or participants who have withdrawn their consent. Latent class analysis of smoking was carried out using Mplus (version 8.8)^31^ via the MplusAutomation R package^32^. All downstream analysis were conducted in R (version 4.2.2)^33^ (Supplementary Text 5).

### Statistical analysis

Longitudinal patterns of smoking were based on 12 repeated 3-category measures of smoking collected from 13 to 28 years. These were derived using longitudinal latent class analysis (LLCA). Patterns using a 4-category smoking variable, and latent class growth analysis, were also derived however these were not used for the association analyses. The associations between “most likely” latent class assignments and each risk factor measure were assessed using a three-step approach. Further details on this and how the best model was selected can be found in Supplementary Text 3.

Univariable logistic regression was used to assess associations across all pairwise comparisons between the derived latent classes of smoking. Odds ratios (ORs) and confidence intervals (CIs) were used to determine the strength of evidence and direction of association. Only results from tests where the 95% CIs did not cross the null are discussed here, however all results can be found in Supplementary Table S16. Multivariable analysis was not carried out given the exploratory nature of the study and number of tests run, each of which will require different covariates to be considered and thus was not logistically feasible.

## Results

### Smoking prevalence

The proportion of participants who reported smoking, non-smoking, or were missing is shown in Figure 1A. Regular and occasional smoking increased in adolescence and early adulthood, then decreased from 24 years. Non-smoking decreased in adolescence but increased in early adulthood. Missingness peaked from 18-21 years.

**Figure 1.**
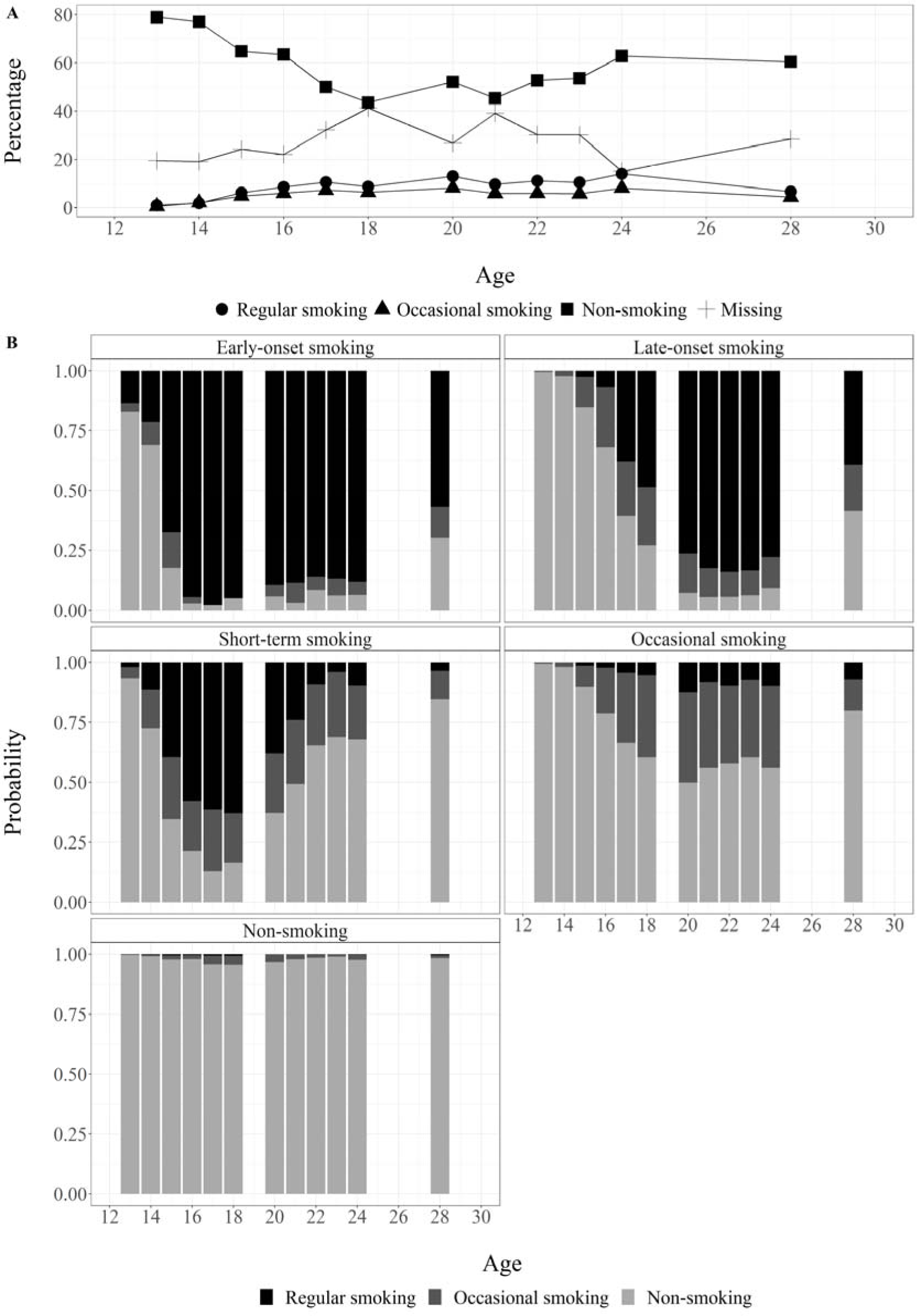
(A) Self-reported smoking, non-smoking and missingness by timepoint (B) probability of regular, occasional, and non-smoking in each of 5 latent classes derived using longitudinal latent class analysis.

### Longitudinal patterns of smoking

A five-class solution using LLCA best explained the relationship between repeated measures of smoking. These five classes (Figure 1B) are described as (1) early-onset smoking, (2) late-onset smoking, (3) occasional smoking, (4) short-term smoking, and (5) non-smoking. Non-smoking was the largest class (62%) and occasional smoking the second largest (14%). There were slightly more participants assigned to late-onset smoking (10%) compared to early-onset smoking (8.1%). Short-term smoking was the smallest class (5.4%).

The early and late-onset smoking classes show a large proportion of regular smoking. The late-onset and short-term smoking classes appear similar in adolescence, but participants assigned to the short-term smoking class show increasing proportions of non-smoking in adulthood. Collectively the early-onset, late-onset and short-term smoking classes are referred to as regular smoking. Early-onset and late-onset smoking are jointly referred to as sustained smoking.

### Associations with risk factors

Summary statistics describing the associations of the 402 included risk factor measures, across 10 pairwise comparisons (5 latent classes of smoking) are visualised using circle plots (Supplementary Figures S10) and interactive Miami plots: https://alexandrayas.github.io/interactive_plts_longitudinal_smoking_associations.html

Of the included risk factor measures 358 (89%) showed evidence of an association in at least one comparison. Two risk factors, cannabis use (16y) and peer smoking (16y, 18y), were associated with smoking patterns in all 10 comparisons. 44 measures did not show evidence of any association. Table 1 shows the number of associated measures in each comparison.

**Table 1.** Number of associated risk factor measures by comparison.

|  |  | Comparison |  |  |  |
| --- | --- | --- | --- | --- | --- |
|  |  | Early-onset smoking | Late-onset smoking | Short-term smoking | Occasional smoking |
| Reference | Non-smoking | 288 (72%*) | 237 (59%) | 183 (46%) | 141 (35%) |
|  | Occasional smoking | 264 (66%) | 162 (40%) | 170 (42%) |  |
|  | Short-term smoking | 125 (31%) | 85 (21%) |  |  |
|  | Late-onset smoking | 161 (40%) |  |  |  |

\* Number of associated measures / 402 (the number of measures included in total) \* 100

Tables 2 and 3 show the top five risk factor measures with the largest ORs in each pairwise comparison. Similar tables showing associations with the smallest CIs or largest population attributable fractions (PAF) or excluding measures with smaller sample sizes (n < 1%) can be found in Supplementary Tables S18-S22. General themes and findings are discussed below.

**Table 2.** Top 5 preceding risk factors.

| **Ref.** | **Comp.** | **Risk factor measure** | **OR** | **LCI** | **UCI** |
| --- | --- | --- | --- | --- | --- |
| **Non-smoking** | **Early-onset smoking** | Mother cannabis use, before pregnancy (0y) | 6.23 | 4.16 | 9.31 |
|  |  | Mother cannabis use, last 2 years (9y) | 5.92 | 3.76 | 9.33 |
|  |  | Mother cannabis use, since child 5 years old (6y) | 5.5 | 3.57 | 8.46 |
|  |  | Mother cannabis use, since child 8 months old | 4.94 | 3.00 | 8.15 |
|  |  | Mother cannabis use, since child 18 months old | 4.87 | 3.05 | 7.79 |
|  | **Late-onset smoking** | Mother cannabis use, last 2 years (9y) | 3.54 | 2.22 | 5.65 |
|  |  | Mother cannabis use, past year (4y) | 2.99 | 1.84 | 4.88 |
|  |  | Mother cannabis use, since child 8 months old | 2.94 | 1.74 | 4.96 |
|  |  | Mother cannabis use, before pregnancy (0y) | 2.92 | 1.88 | 4.54 |
|  |  | Mother cannabis use, since child 5 years old (6y) | 2.61 | 1.61 | 4.23 |
|  | **Occasional smoking** | Mother cannabis use, last 2 years (9y) | 3.54 | 2.00 | 6.26 |
|  |  | Sex assigned at birth (0y) | 3.28 | 2.40 | 4.49 |
|  |  | Mother cannabis use, since child 18 months old | 2.56 | 1.36 | 4.80 |
|  |  | Mother cannabis use, past year (4y) | 2.51 | 1.34 | 4.71 |
|  |  | Ever drank alcohol without permission (8y) | 2.48 | 1.40 | 4.38 |
|  | **Short-term smoking** | Mother cannabis use, last 2 years (9y) | 3.1 | 2.01 | 4.77 |
|  |  | Mother cannabis use, since child 8 months old | 2.86 | 1.79 | 4.59 |
|  |  | Mother cannabis use, since child 18 months old | 2.75 | 1.76 | 4.28 |
|  |  | Mother cannabis use, past year (4y) | 2.4 | 1.51 | 3.81 |
|  |  | Mother cannabis use, past year (5y) | 2.35 | 1.52 | 3.63 |
| **Occasional smoking** | **Early-onset smoking** | Mother smoked last 2m pregnancy (8w) | 4.98 | 3.48 | 7.15 |
|  |  | Mother smokes daily (8w) | 4.6 | 3.31 | 6.39 |
|  |  | Mother smoked last 2w pregnancy (0y) | 4.49 | 3.13 | 6.43 |
|  |  | Mother's partner smokes daily (6y) | 4.5 | 3.21 | 6.29 |
|  |  | Mother regularly smoked (8w) | 4.33 | 3.13 | 5.98 |
|  | **Late-onset smoking** | Mother smoked last 2w pregnancy (0y) | 2.54 | 1.77 | 3.63 |
|  |  | Mother smoked last 2m pregnancy (8w) | 2.44 | 1.69 | 3.52 |
|  |  | Mother felt depressed past month (2y) | 2.44 | 1.88 | 3.18 |
|  |  | Mother's partner smokes daily (2y) | 2.42 | 1.77 | 3.31 |
|  |  | Household members smoke (3y) | 2.38 | 1.81 | 3.13 |
|  | **Short-term smoking** | Sex assigned at birth (0y) | 3.69 | 2.63 | 5.18 |
| | | Mother cotinine $\geq 50$ng/ml first trimester (0y) | 3.57 | 1.84 | 6.90 |
|  |  | Mother's partner smokes daily (6y) | 2.89 | 2.00 | 4.17 |
|  |  | Mother's partner smokes daily (9y) | 2.75 | 1.88 | 4.02 |
|  |  | Ever drank alcohol without permission (8y) | 2.52 | 1.23 | 5.13 |
| **Short-term smoking** | **Early-onset smoking** | Mother cannabis use before pregnancy (0y) | 4.55 | 2.18 | 9.51 |
|  |  | Mother cannabis use, since child 5 years old (6y) | 3.78 | 1.74 | 8.22 |
|  |  | Mother's partner economic activity (0y) | 3.64 | 1.71 | 7.75 |
|  |  | Mother cannabis use, past year (5y) | 3.24 | 1.47 | 7.14 |
|  |  | Maternal grandfather alcohol problem (0y) | 2.86 | 1.33 | 6.14 |
|  | **Late-onset smoking** | Sex assigned at birth (0y) | 0.21 | 0.15 | 0.30 |
|  |  | Maternal grandfather alcohol problem (8y) | 2.64 | 1.19 | 5.88 |
|  |  | Maternal grandfather mental health problem (8y) | 2.09 | 1.16 | 3.77 |
| | | Mother cotinine $\geq 50$ng/ml first trimester (0y) | 0.49 | 0.24 | 0.97 |
|  |  | Part of clubs or plays sports (11y) | 0.61 | 0.44 | 0.84 |
| **Late-onset smoking** | **Early-onset smoking** | Friends smoke cigarettes (10y) | 2.96 | 1.78 | 4.91 |
|  |  | Sex assigned at birth (0y) | 2.56 | 1.93 | 3.40 |
|  |  | Mother cannabis use, since child 18 months old | 2.49 | 1.34 | 4.61 |
| | | Mother cotinine $\geq 50$ng/ml first trimester (0y) | 2.47 | 1.30 | 4.69 |
|  |  | Mother smokes daily (7y) | 2.42 | 1.74 | 3.35 |

**Table 3.** Top 5 concurrent risk factors.

| Ref. | Comp. | Risk factor measure | OR | LCI | UCI |
| --- | --- | --- | --- | --- | --- |
| Non-smoking | Early-onset smoking | Ever use cannabis (17y) | 74.8 | 49.2 | 113.8 |
|  |  | Ever use cannabis (18y) | 61.8 | 37.0 | 103.1 |
|  |  | Ever use cannabis (16y) | 50.4 | 35.7 | 71.30 |
|  |  | Ever use cannabis (18y) | 44.6 | 26.9 | 74.03 |
|  |  | Friends smoke cigarettes (18y) | 32.4 | 21.6 | 48.50 |
|  | Late-onset smoking | Ever use cannabis (20y) | 12.6 | 9.30 | 17.08 |
|  |  | Ever use cannabis (18y) | 11.7 | 8.68 | 15.83 |
|  |  | Ever drug use (20y) | 11.6 | 8.86 | 15.21 |
|  |  | Ever use cannabis (24y) | 11.2 | 8.01 | 15.89 |
|  |  | Ever use cannabis (22y) | 10.9 | 8.04 | 15.01 |
|  | Occasional smoking | Ever use cannabis (18y) | 61.3 | 34.7 | 108.4 |
|  |  | Ever use cannabis (17y) | 44.1 | 30.0 | 64.94 |
|  |  | Ever use cannabis (18y) | 37.0 | 22.2 | 61.76 |
|  |  | Ever use cannabis (16y) | 28.9 | 20.9 | 40.11 |
|  |  | Ever use cannabis (14y) | 22.9 | 15.8 | 33.25 |
|  | Short-term smoking | Ever use cannabis (24y) | 22.4 | 15.6 | 32.19 |
|  |  | Ever use cannabis (22y) | 14.0 | 10.7 | 18.35 |
|  |  | Ever use cannabis (20y) | 11.4 | 9.01 | 14.45 |
|  |  | Ever drug use (24y) | 11.2 | 8.63 | 14.64 |
|  |  | Ever drug use (22y) | 7.80 | 6.29 | 9.67 |
| Occasional smoking | Early-onset smoking | Ever use cannabis (16y) | 16.2 | 11.2 | 23.4 |
|  |  | Friends smoke cigarettes (16y) | 16.0 | 11.3 | 22.87 |
|  |  | Ever use cannabis (17y) | 13.5 | 8.78 | 20.77 |
|  |  | Friends smoke cigarettes (18y) | 11.7 | 7.67 | 18.06 |
|  |  | Ever use cannabis (14y) | 9.67 | 6.39 | 14.66 |
|  | Late-onset smoking | Economic activity (23y) | 3.79 | 2.07 | 6.95 |
|  |  | Mother currently smokes (22y) | 3.64 | 2.10 | 6.32 |
|  |  | Mother smokes daily (18y) | 3.52 | 2.03 | 6.11 |
|  |  | Friends get drunk (20y) | 0.33 | 0.17 | 0.64 |
|  |  | Mother currently smokes (18y) | 2.97 | 1.79 | 4.92 |
|  | Short-term smoking | Friends smoke cigarettes (16y) | 9.99 | 7.07 | 14.12 |
|  |  | Ever use cannabis (16y) | 9.3 | 6.57 | 13.18 |
|  |  | Ever use cannabis (14y) | 8.59 | 5.61 | 13.13 |
|  |  | Ever use cannabis (18y) | 8.56 | 4.77 | 15.39 |
|  |  | Ever use cannabis (17y) | 7.96 | 5.34 | 11.86 |
| Short-term smoking | Early-onset smoking | DAWBA: Lost interest (16y) | 3.18 | 1.39 | 7.29 |
|  |  | CIS-R: Moderate depression (18y) | 2.85 | 1.52 | 5.37 |
|  |  | Mother economic activity (18y) | 0.37 | 0.19 | 0.72 |
|  |  | Friends drink alcohol (16y) | 2.52 | 1.32 | 4.81 |
|  |  | Cannabis use, past year (28y) | 2.46 | 1.61 | 3.75 |
|  | Late-onset smoking | Ever use cannabis (18y) | 0.12 | 0.07 | 0.23 |
|  |  | Ever use cannabis (17y) | 0.13 | 0.09 | 0.20 |
|  |  | Ever use cannabis (14y) | 0.14 | 0.09 | 0.22 |
|  |  | Friends smoke cigarettes (16y) | 0.14 | 0.10 | 0.21 |
|  |  | Ever use cannabis (16y) | 0.15 | 0.10 | 0.21 |
| Late onset smoking | Early-onset smoking | Ever use cannabis (17y) | 12.9 | 8.26 | 20.28 |
|  |  | Ever use cannabis (16y) | 11.7 | 8.06 | 17.16 |
|  |  | Friends smoke cigarettes (16y) | 11.2 | 7.80 | 16.13 |
|  |  | Friends use drugs (16y) | 8.40 | 5.05 | 13.98 |
|  |  | Ever use cannabis (14y) | 8.29 | 5.26 | 13.06 |

### Family and peer substance use

Maternal ever smoking, as well as peer smoking, increased the odds of any smoking versus non-smoking, regular smoking versus occasional smoking, and early-onset smoking. Maternal smoking during childhood and pregnancy also increased the odds of regular smoking versus non-smoking, but showed weaker associations with occasional smoking versus non-smoking. Participants where the mother reported quitting smoking showed lower odds of regular smoking compared to non-smoking and occasional smoking, and early-onset compared to short-term and late-onset smoking. Maternal smoking in adulthood, paternal smoking, and household smoking increased the odds of regular smoking and early-onset smoking compared to less harmful smoking patterns, but decreased the odds of occasional smoking versus non-smoking. Maternal ever smoking at age 18, but not at age 22, decreased the odds of early-onset versus late-onset smoking. Peer smoking decreased the odds of late-onset smoking versus short-term smoking. Smoking by the maternal grandparents showed weaker associations but there was some evidence that this also increased the odds of more harmful smoking patterns.

Familial smoking was more strongly associated with early-onset smoking than other smoking patterns. There were stronger associations with peer smoking than with parental or household smoking. Familial smoking did little to differentiate late-onset from short-term smoking patterns but peer smoking did. The magnitude of associations between peer smoking and longitudinal smoking patterns increased over time during adolescence in all comparisons other than when comparing early-onset to short-term smoking. These associations attenuated towards the null in adulthood.

Greater maternal alcohol consumption increased the odds of any smoking compared to non-smoking, and late-onset smoking compared to short-term smoking. However participants with mothers who drank alcohol more regularly showed lower odds of short-term smoking compared to occasional smoking, and early-onset smoking compared to occasional and late-onset smoking. Paternal alcohol consumption showed similar associations with longitudinal smoking patterns, but did not appear to differentiate early-onset from late-onset smoking. The magnitude of associations between familial alcohol consumption and smoking were slightly stronger for occasional and late-onset smoking than others patterns. Maternal cannabis use increased the odds of any smoking versus non-smoking, and early-onset smoking versus all other smoking patterns.

Peer substance use, including alcohol, cannabis and other drugs, increased the odds of any smoking versus non-smoking, and early-onset smoking versus all other smoking patterns. Peer substance use decreased the odds of late-onset versus short-term smoking. During adolescence peer substance use increased the odds of early-onset and short-term smoking, but not late-onset smoking, compared to occasional smoking. Peer alcohol consumption at age 20 reduced the odds, while peer cannabis use at age 20y increased the odds, of regular versus occasional smoking. The magnitude of associations between peer substance use and smoking patterns increased over time during adolescence, but attenuated in the direction of the null into adulthood.

### Family sociodemographic factors

Lower maternal educational attainment at birth increased the odds of early-onset smoking compared to all other smoking patterns, and short-term smoking versus occasional smoking. Lower paternal educational attainment at birth increased odds of early-onset smoking versus non-smoking, and regular smoking versus occasional smoking. Participants whose father, or maternal grandparents, had lower educational attainment showed lower odds of occasional smoking versus non-smoking. Paternal education was more strongly associated than maternal education with regular versus occasional smoking, however only maternal education showed an association with sustained versus short-term smoking, and early- versus late-onset smoking.

Lower household income at age 18 increased the odds of regular smoking versus non-smoking and occasional smoking. Lower household income at age 11 increased the odds of early-onset compared to non-smoking, occasional smoking, and late-onset smoking. Parental economic inactivity at birth increased the odds of early-onset smoking compared to all other smoking patterns. Paternal economic inactivity at birth, but not later measures, increased the odds of occasional smoking versus non-smoking, but decreased the odds of short-term compared to occasional smoking. Paternal economic inactivity at age 18, but not age 22, increased the odds of short-term smoking versus non-smoking. Maternal economic inactivity at age 8 lowered the odds of late-onset smoking versus non-smoking and occasional smoking. Maternal economic inactivity at age 18 lowered the odds of sustained smoking compared to non-smoking and short-term smoking.

Participants where mothers were tenants or unmarried showed higher odds of early-onset smoking compared to all other smoking patterns, and regular smoking compared to non-smoking and occasional smoking. Measures at birth, but not later, also increased the odds of late-onset versus short-term smoking. Participants with mothers who rented showed lowered odds of occasional versus non-smoking. The association of marital status with sustained versus short-term smoking gradually attenuated towards to null from ages 7 to 22 where participants with unmarried mothers at 22 years had lower odds of late-onset versus short-term smoking.

Participants whose parents reported lower occupational class at birth showed increased odds of short-term versus occasional smoking. Lower maternal occupational class increased the odds of late-onset versus occasional smoking. Lower paternal occupational class increased the odds of early-onset smoking compared to non-smoking, occasional smoking, and late-onset smoking.

Maternal neighbourhood deprivation increased the odds of regular smoking compared to non-smoking and occasional smoking, and sustained compared to short-term smoking. Participants whose mother lived in more rural areas showed lower odds of early-onset smoking compared to occasional and late-onset smoking.

Family sociodemographic factors were more strongly associated with sustained smoking, particularly early-onset smoking, than other smoking patterns. Maternal home ownership and marital status were more strongly associated with smoking patterns than other family sociodemographic factors such as neighbourhood deprivation.

### Individual lifestyle factors

Participants with a higher than median BMI showed increased odds of regular smoking compared to non-smoking and occasional smoking. These associations were slightly stronger when investigating short-term smoking compared to other patterns. Having a higher BMI at some later timepoints lowered the odds of occasional smoking versus non-smoking (14, 24y), sustained versus short-term smoking (16, 18y), and early- versus late-onset smoking (18y).

Higher calorie intake at age 7 lowered the odds of late-onset smoking compared to non-smoking and short-term smoking. At age 14 higher calorie intake lowered the odds of any smoking compared to non-smoking, but increased the odds of late-onset versus short-term smoking. Eating less healthy food at age 14 increased the odds of early-onset smoking compared to non-smoking. However less healthy diets at age 14 decreased the odds of occasional smoking compared to non-smoking, and early-onset and short-term smoking compared to occasional smoking.

Fewer days of moderate-to-vigorous physical activity (MVPA) at ages 11 and 14 increased the odds of any smoking compared to non-smoking, and early-onset and short-term smoking compared to occasional smoking. Lower MVPA at age 14 also increased the odds of early-onset compared to late-onset smoking, but lowered the odds of late-onset versus short-term smoking. Lower MVPA at age 16 increased the odds of regular smoking compared to non-smoking and occasional smoking. Not playing sports at 11 years increased the odds of short-term smoking compared to non-smoking and occasional smoking. However this decreased the odds of late-onset and occasional smoking compared to non-smoking, and sustained compared short-term smoking. At age 14 not playing sports increased the odds early-onset smoking versus all other smoking patterns, but decreased the odds of late-onset smoking compared to non-smoking and short-term smoking.

Spending fewer hours asleep increased the odds of late-onset smoking versus non-smoking. At age 11 less sleep decreased the odds of early- versus late-onset smoking. At age 16 less sleep increased the odds of early-onset and short-term smoking compared to both non-smoking and occasional smoking, and early-onset compared to late-onset smoking. At age 25 less sleep increased the odds of occasional smoking versus non-smoking, and sustained versus short-term smoking, but decreased the odds of short-term versus occasional smoking.

Other substance use, including greater alcohol consumption, cannabis use, and other drug use, increased the odds of any smoking versus non-smoking. The associations of other substance use with smoking patterns peaked at around 16 years of age for alcohol, 17 years for cannabis use, and 18 years for other drug use. Other substance use during adolescence also increased the odds of early-onset versus late-onset smoking, but decreased the odds of late-onset compared to short-term smoking.

Alcohol consumption during adolescence increased the odds of regular compared to occasional smoking. Later adulthood measures of alcohol consumption showed weaker associations and the opposite direction of association as observed in adolescence. In adulthood more regular alcohol consumption decreased the odds of early-onset and short-term smoking compared to occasional smoking, and early- versus late-onset smoking, but increased the odds of late-onset versus short-term smoking. Later measures of cannabis and other drug use in adulthood decreased the odds of short-term smoking versus occasional smoking, but increased odds of sustained versus short-term smoking. The association of other substance use with smoking patterns attenuated from adolescence into adulthood.

Other substance use was more strongly associated with smoking patterns than other lifestyle factors, and there was stronger evidence of associations with early-onset and short-term smoking than other smoking patterns.

### Individual sociodemographic factors

Participants assigned female at birth showed greater odds of early-onset and short-term smoking compared to non-smoking and occasional smoking, and early-onset compared to late-onset smoking. However, female participants showed lower odds of late-onset versus occasional smoking, and sustained versus short-term smoking.

Participants who planned to leave education at any point (14, 16, 18y) showed increased odds early-onset smoking compared to non-smoking, occasional, and late-onset smoking, and short-term versus occasional smoking. Participants planning to leave education after Year 11 (16y) showed increased odds of any smoking versus non-smoking, but decreased odds of late-onset versus short-term smoking. Participants who did not plan to attend university showed increased odds of short-term smoking versus non-smoking, but decreased odds of late-onset versus short-term smoking. Planning to leave education after Year 11 (14y), and not planning to attend university (18y), decreased the odds of occasional smoking versus non-smoking. In the UK education system, Year 11 refers to the eleventh year of compulsory education, when students are usually around 15 to 16 years old and preparing for their General Certificate of Secondary Education (GCSE) examinations.

Participants currently studying non-A level qualifications (18y) had lower odds of early-onset smoking compared to non-smoking, occasional smoking, and late-onset smoking, and short-term smoking compared to non-smoking and occasional smoking. “Advanced Level” (A-level) qualifications typically follow GCSEs or equivalent, usually in the age range of 16 to 18 years. Lower educational attainment at ages 18 and 20 increased the odds of regular smoking and early-onset smoking, but decreased the odds of late-onset versus short-term smoking. Lower educational attainment at age 20 also lowered the odds of occasional smoking versus non-smoking. Later measures of education should participants who did not go on to higher education showed increased odds of early-onset smoking compared to non-smoking, occasional smoking and late-onset smoking, and short-term smoking versus non-smoking and occasional smoking. However this lowered the odds of late-onset versus short-term smoking.

Participants who were not economically active, or in education or training, had higher odds of regular smoking compared to non-smoking and occasional smoking. Participants who were not in education or training also showed increased odds of early-onset compared to late-onset smoking. Participants who earned a higher income (25y) showed lower odds of regular smoking compared to non-smoking and occasional smoking, and early-onset smoking compared to short-term smoking. However higher income increased the odds of occasional smoking versus non-smoking. Unemployment at age 25 increased the odds of early-onset and occasional smoking versus non-smoking. Shift and night work increased the odds of early-onset smoking compared to non-smoking, short-term smoking and late-onset smoking, and occasional smoking versus non-smoking. However, shift and night work both lowered the odds of late-onset versus occasional smoking. Night shifts also lowered the odds of short-term versus occasional smoking. Lower social class increased the odds of regular smoking compared to non-smoking and occasional smoking.

Neighbourhood deprivation increased the odds of sustained smoking versus non-smoking and occasional smoking, and early-onset smoking compared to all other smoking patterns. Living rurally, particularly at age 18, decreased the odds of early-onset smoking compared to non-smoking, occasional smoking and late-onset smoking, and short-term smoking compared to non-smoking and occasional smoking. Rural living at age 18 also increased the odds of late-onset versus short-term smoking.

Factors related to education were more strongly associated with smoking than other sociodemographic factors and showed stronger associations with early-onset smoking compared to other smoking patterns.

### Mental health and other factors

Familial mental health problems increased the odds of regular smoking versus non-smoking and occasional smoking. Familial mental health did not appear to associate with sustained versus short-term smoking, and only a few measures increased the odds of early-onset versus late-onset smoking. Participants who reported having mental health problems or lower wellbeing showed increased odds of any smoking versus non-smoking, and early-onset smoking compared to all other smoking patterns. There were fewer associations between mental health and short-term or late-onset smoking. However having chronic fatigue (18y) increased the odds of short-term versus occasional smoking, and decreased the odds of late-onset versus short-term smoking. Lower wellbeing at age 18 increased the odds of short-term versus occasional smoking. Lower wellbeing at age 23 increased the odds of late-onset smoking versus short-term smoking. Lower wellbeing at age 23, and reporting a mental health problem at age 22, also increased the odds of late-onset versus occasional smoking.

Participants with more adverse childhood experiences (ACEs) or trauma showed greater odds of any smoking versus non-smoking, and regular smoking versus occasional smoking. Trauma also increased the odds of early-onset smoking compared to short-term smoking and late-onset smoking. The extended ACEs score (0-16y) suggested participants with 5 or more ACEs had lower odds of late-onset compared to short-term smoking.

Participants who had become a parent in their twenties showed increased odds of regular smoking compared to non-smoking and occasional smoking, and early-onset compared to late-onset smoking. However parenthood decreased the odds of late-onset versus short-term smoking. Participants who had ever been pregnant at age 21 or reported being a parent at age 22, but not later, showed increased odds of early-onset smoking compared to short-term smoking. Parenthood at age 28 reduced the odds of occasional smoking versus non-smoking.

## Discussion

This study aimed to synthesise the association of 402 risk factor measures with five latent classes of smoking, derived using 12 repeated smoking measures collected from 13 to 28 years of age. Parental and peer substance use, parental SEP, other substance use, physical activity, education, mental health, ACEs, and parenthood emerged as strong correlates of longitudinal patterns of smoking. The associations of identified risk factors varied depending on the smoking patterns being investigated. When examining any smoking there were differences between regular and occasional smoking patterns in their relationship with familial factors and BMI. In comparisons to short-term smoking, there many differences between early-onset and late-onset smoking patterns in terms of their association with peer and other substance use. Often risk factors were more weakly associated with late-onset smoking than early-onset smoking, and few risk factors differentiated late-onset from short-term smoking.

Findings in this study reiterates that parents, friends, and household smoking^34–37^ influence smoking by many mechanisms^38–41^. While parents influence smoking initiation, they may have less of an effect on smoking cessation. Participants who had become parents in their twenties were also more likely to have smoked in adolescence. Better smoking cessation programs could then be tailored towards expectant parents. These could: (a) raise awareness of parental smoking influences on children’s smoking habits, (b) integrate smoking and substance use prevention into prenatal and postnatal care services, (c) provide resources, support, and specialized programs for parents with a history of substance use, and (d) increase support for young individuals becoming parents.

Smoking interventions should also emphasise the strong role of peers in smoking behaviours, potentially by establishing peer-led support or mentoring groups and encouraging positive peer norms through school-based initiatives. These could (a) integrate substance use prevention into the curricula, (b) implement comprehensive education on the risks of various substances, (c) provide counselling and support for pupils with a history of substance use, and (d) increase support for students from lower socio-economic backgrounds or students not going on to further education and (e) introduce smoking prevention components into vocational training programs.

Smoking occurs disproportionately more in disadvantaged groups and the role of parental SEP^42^ and education^43,44^ in smoking has been shown. While this may be protective against smoking this occurs less in marginalized groups^45^. In this study parental sociodemographic factors did not consistently associate with all smoking patterns when compared to non-smoking. Untargeted smoking cessation campaigns in Europe may exacerbate health inequalities^46^ so future studies should consider intersections between different risk factors. Engaging communities in the development and implementation of smoking prevention initiatives may also improve their effectiveness, and could communicate the role of neighbourhood deprivation in smoking, and establish community partnerships to address social determinants in smoking. Local governments could also (a) develop more policies to address smoking disparities related to socioeconomic position, (b) implement financial support programs for low-income families, and (c) increase smoke-free public spaces in deprived areas. Smoking prevention campaigns tailored to individuals who are economically inactive or doing shift or night work could also reduce smoking prevalence. These individuals should be provided with support and resources. Collaboration with employers to create smoke-free workplaces and implement workplace smoking cessation incentives may also be effective.

Participants who use cannabis or other drugs are likely to smoke^47^. Although drug use is often underreported, it does not occur in a large proportion of the adult UK population and these behaviours change rapidly over time. This can obscure statistics particularly when an outcome completely separates a predictor (Supplementary Figures S7-S9). These associations could also be confused for a gateway hypothesis when confounding is not sufficiently handled so future research could control for risk factors identified here. Interventions focused on other risk factors such as exercise, household income, and mental health, which show smaller but more consistent effects, may reduce smoking more at a population-level. This would support other findings of a bidirectional effect between smoking and mental health^48^, and how co-use of cannabis and tobacco is also related with worse mental health^49^. Any smoking prevention or cessation programmes should then integrate mental health support and be trauma-informed. Other work has shown that smoking susceptibility associates with activity and weight^50^, echoing the association of BMI and physical activity in this study. School-based programs could further encourage good nutrition, physical activity, and adequate sleep.

More generally, any smoking prevention or cessation programs should (a) use diverse media channels to reach different audiences, (b) tailor strategies to address different patterns of smoking and stages of life, (c) be accessible and affordable, (d) promote the use of helplines, mobile apps, and online resources for quitting, and (e) implement follow-up programs to assess their long-term effectiveness.

A multifaceted approach that considers individual, family, and community factors is clearly essential for effective smoking prevention and cessation efforts.

### Strengths and limitations

The strengths of this study include the use of rich longitudinal smoking data, large numbers of investigated risk factors, repeated measures, and nuanced pairwise comparisons between all smoking patterns. Limitations include using complete cases rather than multiple imputation meaning attrition may influence findings. Sample sizes varied across comparisons, risk factors, and timepoints, meaning statistics using larger latent classes, or earlier measures, are more well powered. Binary measures were used meaning the effect estimates could depend on the parameterisation or thresholds used. Univariable analysis does not account for confounding or effect modification, nor the interplay between several measures.

### Implications

Many risk factors for initiating smoking may not act in the same way for regular or sustained smoking and vary depending on age of onset. Associations may also vary due to the measurement and age. This suggests tobacco policies and interventions should take a more holistic approach, considering the interconnectedness of many risk factors, but be tailored towards specific smoking patterns rather than tackling smoking broadly.

## Data Availability

Data used in this project and any resulting data from the analyses are available on request to the ALSPAC Executive Committee and subject to a data access fee. The ALSPAC data management plan (http://www.bristol.ac.uk/alspac/researchers/data-access/) describes in detail the policy regarding data sharing, which is through a system of managed open access.

## Data Access and Sharing

Data used in this project and any resulting data from the analyses are available on request to the ALSPAC Executive Committee and subject to a data access fee. The ALSPAC data management plan (http://www.bristol.ac.uk/alspac/researchers/data-access/) describes in detail the policy regarding data sharing, which is through a system of managed open access. The script template used to generate datasets used in this study was dated 13^th^ January 2023.

Ethical approval for the study was obtained from the ALSPAC Law and Ethics committee and local research ethics committees (NHS Haydock REC: 10/H1010/70). Informed consent for the use of data collected via questionnaires and clinics was obtained from participants following the recommendations of the ALSPAC Ethics and Law Committee at the time. Consent for biological samples has been collected in accordance with the Human Tissue Act (2004).

Data access for this project has been granted (B3499) prior to this protocol and the proposed analysis of data was pre-registered on the Open Science Framework (OSF) here: https://osf.io/kp235/.

R scripts used for data analysis are available in a github repository (https://github.com/alexandrayas/ALSPAC_CRUK_smkvap/tree/main/Longitudinal%20patterns%20of%20smoking).

## Author contribution

**AA**: conceptualization; formal analysis; visualization; writing – original draft preparation; writing – review & editing. **LH**: conceptualization; formal analysis; writing – review & editing. **HJ**: conceptualization; formal analysis; writing – review & editing. **JK**: conceptualization; formal analysis; writing – review & editing. **MM**: funding acquisition; supervision; writing – review & editing. **HS**: conceptualization; funding acquisition; supervision; writing – review & editing. **JH**: conceptualization; formal analysis; supervision; writing – review & editing.

## Acknowledgements

We are extremely grateful to all the families who took part in this study, the midwives for their help in recruiting them, and the whole ALSPAC team, which includes interviewers, computer and laboratory technicians, clerical workers, research scientists, volunteers, managers, receptionists, and nurses.

