## Supplementary Figures for "Longitudinal patterns of smoking behaviours in adolescence and early adulthood and their association with modifiable and sociodemographic risk factors"

**Figure S1: Number of risk factor measures included or removed due to sample size**

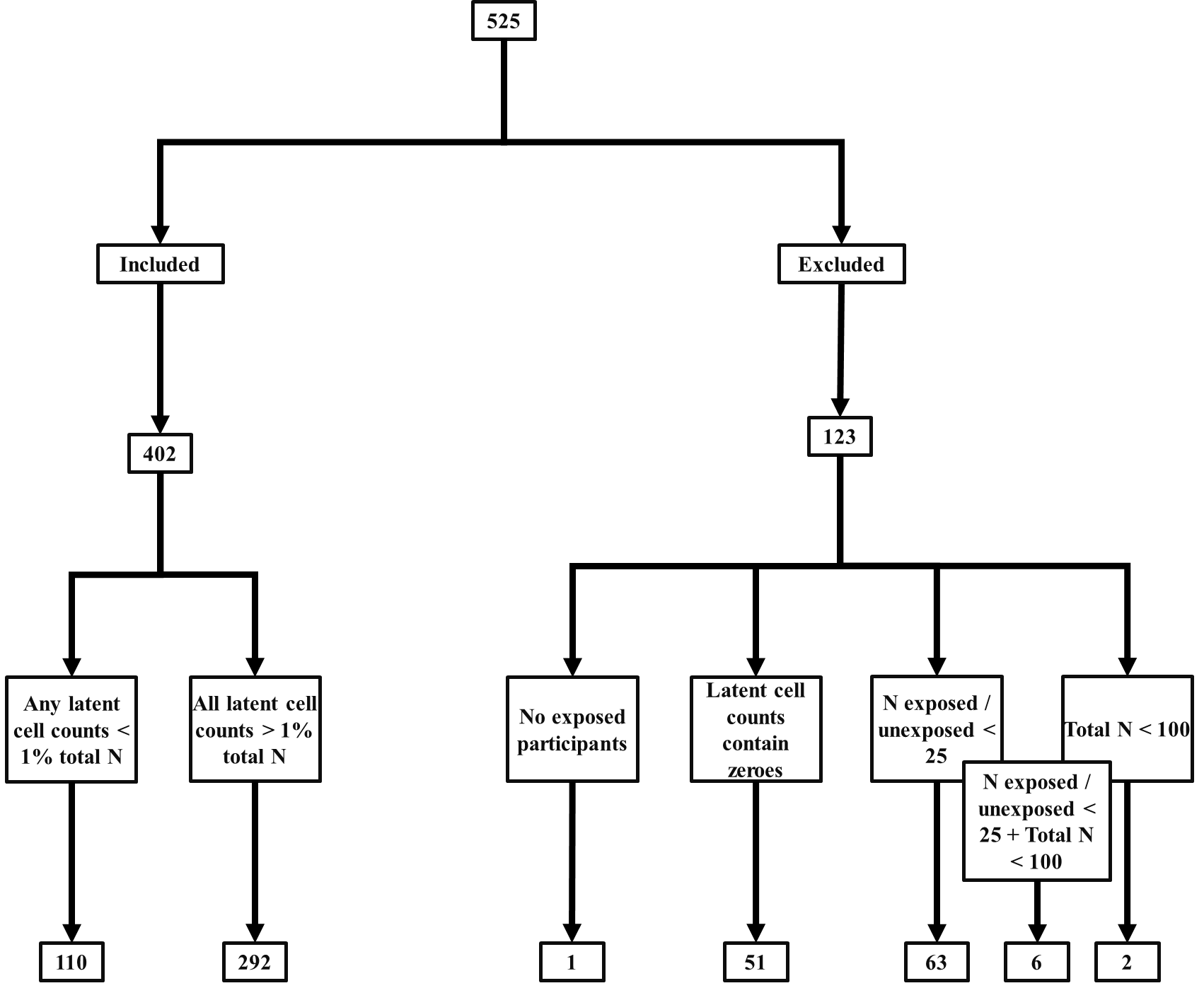

**Figure S2. Graphs showing the probability of each level of smoking frequency for each class over time from the longitudinal latent class model with six latent classes using 4-category smoking measures**

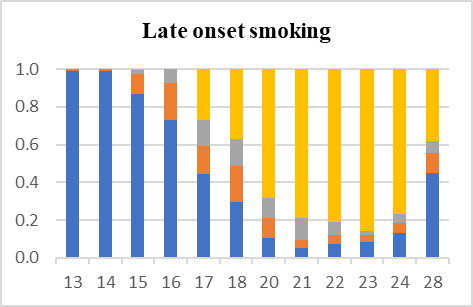

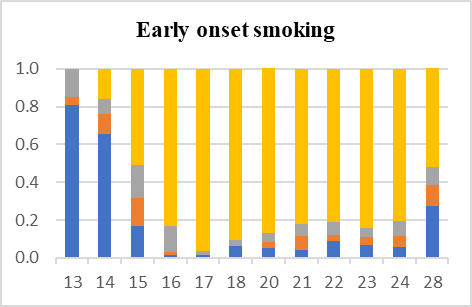

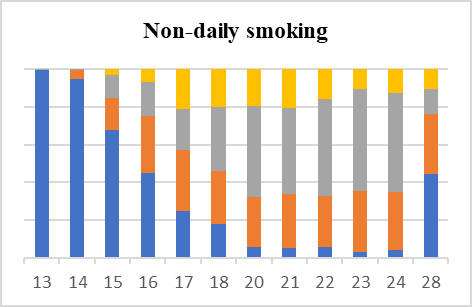

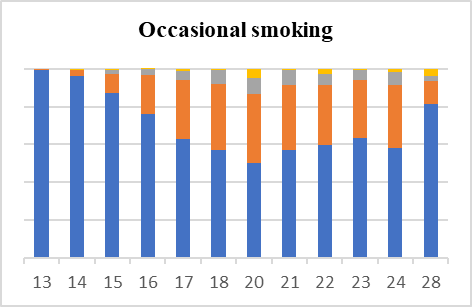

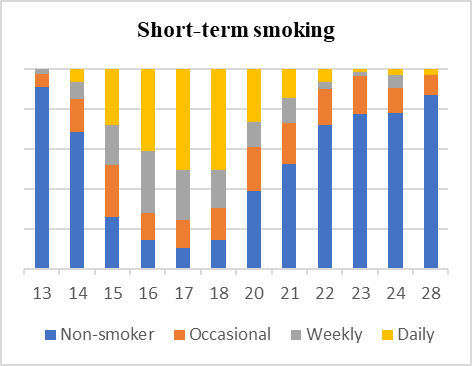

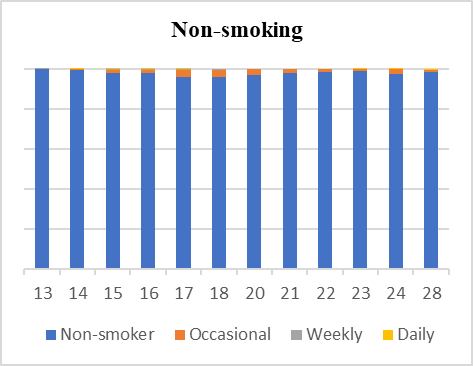

Early onset smoking

|  | Non-smoker | Occasional | Weekly | Daily |
| --- | --- | --- | --- | --- |
| 13 | 0.809 | 0.04 | 0.151 |  |
| 14 | 0.657 | 0.105 | 0.08 | 0.158 |
| 15 | 0.166 | 0.151 | 0.174 | 0.509 |
| 16 | 0.013 | 0.016 | 0.14 | 0.831 |
| 17 | 0.017 | 0 | 0.017 | 0.966 |
| 18 | 0.06 | 0 | 0.035 | 0.904 |
| 20 | 0.053 | 0.033 | 0.047 | 0.868 |
| 21 | 0.041 | 0.074 | 0.065 | 0.82 |
| 22 | 0.087 | 0.036 | 0.068 | 0.809 |
| 23 | 0.067 | 0.041 | 0.048 | 0.844 |
| 24 | 0.059 | 0.058 | 0.076 | 0.806 |
| 28 | 0.272 | 0.115 | 0.091 | 0.523 |

Late onset smoking

|  | Non-smoker | Occasional | Weekly | Daily |
| --- | --- | --- | --- | --- |
| 13 | 0.989 | 0.011 | 0 |  |
| 14 | 0.988 | 0.012 | 0 | 0 |
| 15 | 0.87 | 0.104 | 0.019 | 0.008 |
| 16 | 0.731 | 0.196 | 0.074 | 0 |
| 17 | 0.444 | 0.151 | 0.134 | 0.271 |
| 18 | 0.295 | 0.19 | 0.142 | 0.372 |
| 20 | 0.104 | 0.109 | 0.106 | 0.68 |
| 21 | 0.055 | 0.039 | 0.117 | 0.789 |
| 22 | 0.072 | 0.048 | 0.07 | 0.81 |
| 23 | 0.082 | 0.039 | 0.022 | 0.857 |
| 24 | 0.134 | 0.05 | 0.047 | 0.769 |
| 28 | 0.449 | 0.108 | 0.062 | 0.38 |

Early onset smoking

|  | Non-smoker | Occasional | Weekly | Daily |
| --- | --- | --- | --- | --- |
| 13 | 0.809 | 0.04 | 0.151 |  |
| 14 | 0.657 | 0.105 | 0.08 | 0.158 |
| 15 | 0.166 | 0.151 | 0.174 | 0.509 |
| 16 | 0.013 | 0.016 | 0.14 | 0.831 |
| 17 | 0.017 | 0 | 0.017 | 0.966 |
| 18 | 0.06 | 0 | 0.035 | 0.904 |
| 20 | 0.053 | 0.033 | 0.047 | 0.868 |
| 21 | 0.041 | 0.074 | 0.065 | 0.82 |
| 22 | 0.087 | 0.036 | 0.068 | 0.809 |
| 23 | 0.067 | 0.041 | 0.048 | 0.844 |
| 24 | 0.059 | 0.058 | 0.076 | 0.806 |
| 28 | 0.272 | 0.115 | 0.091 | 0.523 |

Non-daily smoking

|  | Non-smoker | Occasional | Weekly | Daily |
| --- | --- | --- | --- | --- |
| 13 | 0.996 | 0 | 0.004 |  |
| 14 | 0.948 | 0.046 | 0.006 | 0 |
| 15 | 0.675 | 0.171 | 0.122 | 0.032 |
| 16 | 0.452 | 0.3 | 0.178 | 0.07 |
| 17 | 0.249 | 0.321 | 0.217 | 0.213 |
| 18 | 0.182 | 0.277 | 0.339 | 0.202 |
| 20 | 0.058 | 0.267 | 0.481 | 0.194 |
| 21 | 0.051 | 0.288 | 0.453 | 0.208 |
| 22 | 0.056 | 0.271 | 0.513 | 0.161 |
| 23 | 0.03 | 0.323 | 0.541 | 0.107 |
| 24 | 0.044 | 0.303 | 0.524 | 0.129 |
| 28 | 0.443 | 0.317 | 0.134 | 0.106 |

Occasional smoking

|  | Non-smoker | Occasional | Weekly | Daily |
| --- | --- | --- | --- | --- |
| 13 | 0.993 | 0.007 | 0 |  |
| 14 | 0.961 | 0.034 | 0.001 | 0.004 |
| 15 | 0.87 | 0.101 | 0.024 | 0.005 |
| 16 | 0.762 | 0.205 | 0.031 | 0.002 |
| 17 | 0.627 | 0.312 | 0.049 | 0.012 |
| 18 | 0.572 | 0.346 | 0.077 | 0.005 |
| 20 | 0.5 | 0.365 | 0.084 | 0.052 |
| 21 | 0.571 | 0.343 | 0.078 | 0.007 |
| 22 | 0.597 | 0.317 | 0.057 | 0.028 |
| 23 | 0.635 | 0.307 | 0.049 | 0.008 |
| 24 | 0.582 | 0.331 | 0.069 | 0.018 |
| 28 | 0.811 | 0.122 | 0.031 | 0.036 |

Short-term smoking

|  | Non-smoker | Occasional | Weekly | Daily |
| --- | --- | --- | --- | --- |
| 13 | 0.91 | 0.062 | 0.028 |  |
| 14 | 0.685 | 0.164 | 0.086 | 0.065 |
| 15 | 0.259 | 0.261 | 0.197 | 0.283 |
| 16 | 0.144 | 0.134 | 0.309 | 0.412 |
| 17 | 0.105 | 0.14 | 0.25 | 0.504 |
| 18 | 0.146 | 0.157 | 0.191 | 0.507 |
| 20 | 0.39 | 0.219 | 0.126 | 0.265 |
| 21 | 0.522 | 0.209 | 0.124 | 0.145 |
| 22 | 0.72 | 0.18 | 0.032 | 0.069 |
| 23 | 0.772 | 0.192 | 0.02 | 0.016 |
| 24 | 0.777 | 0.125 | 0.067 | 0.032 |
| 28 | 0.869 | 0.098 | 0 | 0.033 |

Non-smoking

|  | Non-smoker | Occasional | Weekly | Daily |
| --- | --- | --- | --- | --- |
| 13 | 0.998 | 0.001 | 0.001 |  |
| 14 | 0.992 | 0.006 | 0.001 | 0.001 |
| 15 | 0.979 | 0.016 | 0.003 | 0.001 |
| 16 | 0.981 | 0.015 | 0.002 | 0.001 |
| 17 | 0.96 | 0.035 | 0.005 | 0.001 |
| 18 | 0.958 | 0.037 | 0.005 | 0 |
| 20 | 0.967 | 0.033 | 0 | 0 |
| 21 | 0.979 | 0.021 | 0 | 0 |
| 22 | 0.985 | 0.015 | 0 | 0 |
| 23 | 0.989 | 0.01 | 0 | 0.001 |
| 24 | 0.976 | 0.022 | 0.001 | 0.001 |
| 28 | 0.983 | 0.012 | 0.001 | 0.003 |

**Figure S3. Graphs showing the probability of each level of smoking frequency for each class over time from the longitudinal latent class model with five latent classes using 3-category smoking measures**

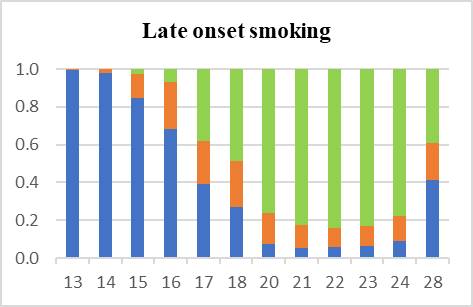

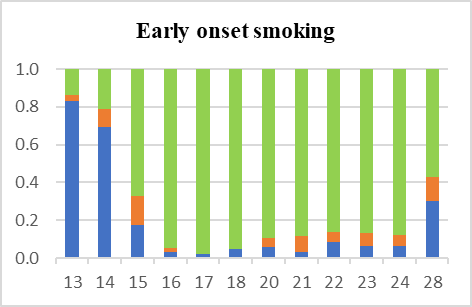

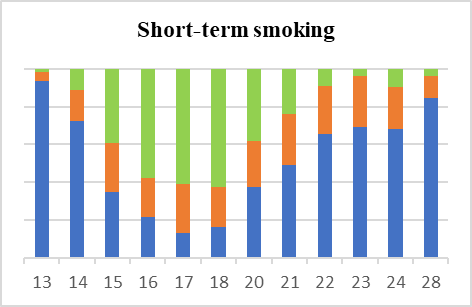

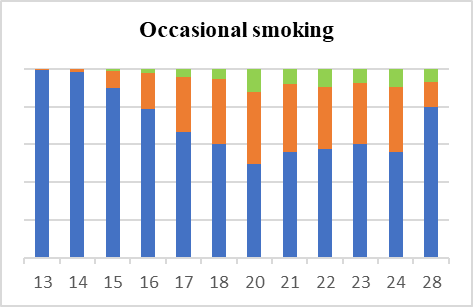

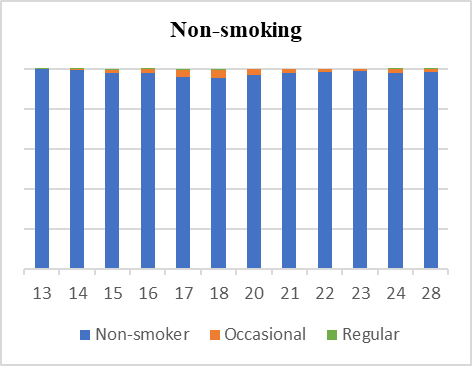

Late onset smoking

|  | Non-smoker | Occasional | Regular |
| --- | --- | --- | --- |
| 13 | 0.995 | 0.005 | 0 |
| 14 | 0.977 | 0.023 | 0 |
| 15 | 0.848 | 0.126 | 0.026 |
| 16 | 0.681 | 0.249 | 0.069 |
| 17 | 0.393 | 0.227 | 0.38 |
| 18 | 0.271 | 0.244 | 0.486 |
| 20 | 0.073 | 0.164 | 0.764 |
| 21 | 0.055 | 0.122 | 0.823 |
| 22 | 0.056 | 0.105 | 0.839 |
| 23 | 0.062 | 0.105 | 0.833 |
| 24 | 0.092 | 0.13 | 0.778 |
| 28 | 0.415 | 0.192 | 0.393 |

Early onset smoking

|  | Non-smoker | Occasional | Regular |
| --- | --- | --- | --- |
| 13 | 0.829 | 0.035 | 0.135 |
| 14 | 0.691 | 0.095 | 0.214 |
| 15 | 0.177 | 0.15 | 0.673 |
| 16 | 0.03 | 0.025 | 0.945 |
| 17 | 0.022 | 0 | 0.978 |
| 18 | 0.05 | 0 | 0.95 |
| 20 | 0.057 | 0.048 | 0.895 |
| 21 | 0.032 | 0.083 | 0.884 |
| 22 | 0.084 | 0.056 | 0.86 |
| 23 | 0.061 | 0.071 | 0.868 |
| 24 | 0.063 | 0.057 | 0.881 |
| 28 | 0.302 | 0.129 | 0.569 |

Short-term smoking

|  | Non-smoker | Occasional | Regular |
| --- | --- | --- | --- |
| 13 | 0.933 | 0.048 | 0.02 |
| 14 | 0.726 | 0.161 | 0.113 |
| 15 | 0.348 | 0.257 | 0.395 |
| 16 | 0.215 | 0.206 | 0.58 |
| 17 | 0.13 | 0.259 | 0.612 |
| 18 | 0.165 | 0.207 | 0.628 |
| 20 | 0.373 | 0.247 | 0.38 |
| 21 | 0.493 | 0.266 | 0.24 |
| 22 | 0.655 | 0.255 | 0.09 |
| 23 | 0.69 | 0.27 | 0.04 |
| 24 | 0.679 | 0.226 | 0.095 |
| 28 | 0.846 | 0.118 | 0.035 |

Occasional smoking

|  | Non-smoker | Occasional | Regular |
| --- | --- | --- | --- |
| 13 | 0.994 | 0.006 | 0 |
| 14 | 0.981 | 0.019 | 0 |
| 15 | 0.9 | 0.087 | 0.014 |
| 16 | 0.786 | 0.191 | 0.023 |
| 17 | 0.665 | 0.293 | 0.043 |
| 18 | 0.604 | 0.343 | 0.054 |
| 20 | 0.498 | 0.378 | 0.124 |
| 21 | 0.562 | 0.356 | 0.083 |
| 22 | 0.578 | 0.326 | 0.096 |
| 23 | 0.604 | 0.322 | 0.074 |
| 24 | 0.562 | 0.341 | 0.096 |
| 28 | 0.799 | 0.129 | 0.072 |

Non-smoking

|  | Non-smoker | Occasional | Regular |
| --- | --- | --- | --- |
| 13 | 0.998 | 0.001 | 0.001 |
| 14 | 0.991 | 0.006 | 0.002 |
| 15 | 0.978 | 0.017 | 0.005 |
| 16 | 0.98 | 0.016 | 0.004 |
| 17 | 0.957 | 0.037 | 0.006 |
| 18 | 0.955 | 0.038 | 0.006 |
| 20 | 0.967 | 0.033 | 0 |
| 21 | 0.979 | 0.021 | 0 |
| 22 | 0.985 | 0.015 | 0 |
| 23 | 0.99 | 0.01 | 0 |
| 24 | 0.977 | 0.022 | 0.002 |
| 28 | 0.984 | 0.012 | 0.004 |

**Figure S4. Graphs showing the probability of each level of smoking frequency for each class over time from the cubic latent class growth model with five latent classes using 3-category smoking measures**

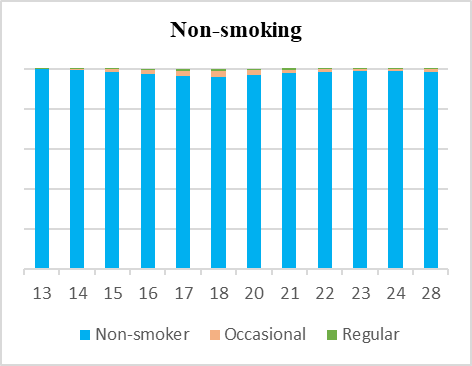

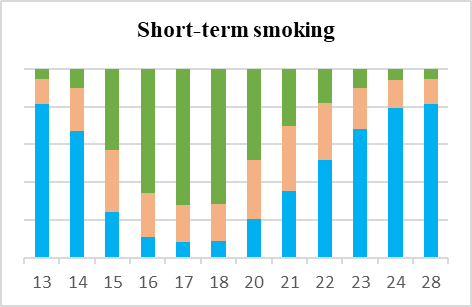

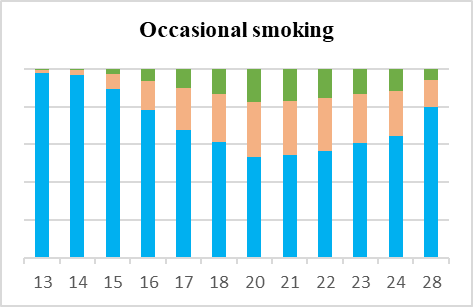

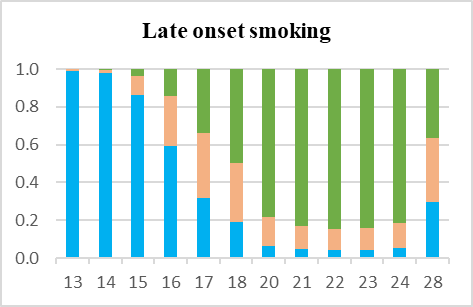

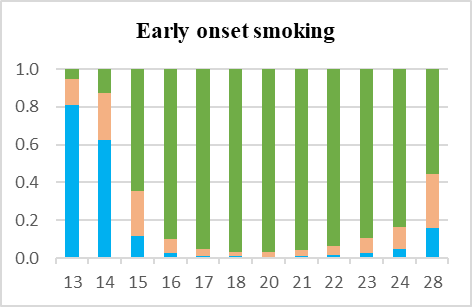

Non-smoking

|  | Non-smoker | Occasional | Regular |
| --- | --- | --- | --- |
| 13 | 0.996 | 0.003 | 0.001 |
| 14 | 0.994 | 0.005 | 0.001 |
| 15 | 0.983 | 0.013 | 0.004 |
| 16 | 0.971 | 0.022 | 0.007 |
| 17 | 0.962 | 0.028 | 0.009 |
| 18 | 0.96 | 0.03 | 0.01 |
| 20 | 0.97 | 0.023 | 0.007 |
| 21 | 0.977 | 0.018 | 0.006 |
| 22 | 0.982 | 0.014 | 0.004 |
| 23 | 0.986 | 0.011 | 0.003 |
| 24 | 0.988 | 0.009 | 0.003 |
| 28 | 0.984 | 0.012 | 0.004 |

Short-term smoking

|  | Non-smoker | Occasional | Regular |
| --- | --- | --- | --- |
| 13 | 0.815 | 0.133 | 0.051 |
| 14 | 0.672 | 0.224 | 0.105 |
| 15 | 0.243 | 0.33 | 0.427 |
| 16 | 0.112 | 0.233 | 0.655 |
| 17 | 0.084 | 0.193 | 0.723 |
| 18 | 0.087 | 0.198 | 0.715 |
| 20 | 0.206 | 0.314 | 0.48 |
| 21 | 0.356 | 0.342 | 0.302 |
| 22 | 0.517 | 0.301 | 0.183 |
| 23 | 0.684 | 0.216 | 0.099 |
| 24 | 0.791 | 0.15 | 0.059 |
| 28 | 0.812 | 0.136 | 0.052 |

Occasional smoking

|  | Non-smoker | Occasional | Regular |
| --- | --- | --- | --- |
| 13 | 0.979 | 0.016 | 0.005 |
| 14 | 0.967 | 0.025 | 0.008 |
| 15 | 0.894 | 0.078 | 0.028 |
| 16 | 0.783 | 0.155 | 0.062 |
| 17 | 0.676 | 0.221 | 0.103 |
| 18 | 0.612 | 0.256 | 0.131 |
| 20 | 0.532 | 0.294 | 0.174 |
| 21 | 0.542 | 0.29 | 0.168 |
| 22 | 0.566 | 0.279 | 0.155 |
| 23 | 0.606 | 0.26 | 0.135 |
| 24 | 0.647 | 0.237 | 0.115 |
| 28 | 0.799 | 0.144 | 0.057 |

Late onset smoking

|  | Non-smoker | Occasional | Regular |
| --- | --- | --- | --- |
| 13 | 0.989 | 0.009 | 0.003 |
| 14 | 0.978 | 0.016 | 0.005 |
| 15 | 0.865 | 0.099 | 0.036 |
| 16 | 0.59 | 0.268 | 0.142 |
| 17 | 0.319 | 0.343 | 0.338 |
| 18 | 0.193 | 0.307 | 0.5 |
| 20 | 0.062 | 0.154 | 0.784 |
| 21 | 0.046 | 0.123 | 0.831 |
| 22 | 0.042 | 0.114 | 0.844 |
| 23 | 0.044 | 0.117 | 0.839 |
| 24 | 0.051 | 0.133 | 0.816 |
| 28 | 0.295 | 0.341 | 0.364 |

Early onset smoking

|  | Non-smoker | Occasional | Regular |
| --- | --- | --- | --- |
| 13 | 0.811 | 0.136 | 0.053 |
| 14 | 0.622 | 0.251 | 0.127 |
| 15 | 0.117 | 0.24 | 0.643 |
| 16 | 0.027 | 0.076 | 0.897 |
| 17 | 0.011 | 0.034 | 0.955 |
| 18 | 0.008 | 0.023 | 0.969 |
| 20 | 0.007 | 0.023 | 0.97 |
| 21 | 0.011 | 0.032 | 0.957 |
| 22 | 0.016 | 0.048 | 0.936 |
| 23 | 0.028 | 0.079 | 0.894 |
| 24 | 0.045 | 0.12 | 0.835 |
| 28 | 0.159 | 0.283 | 0.558 |

**Figure S5. Sample sizes for each risk factor by the proportion of participants exposed**

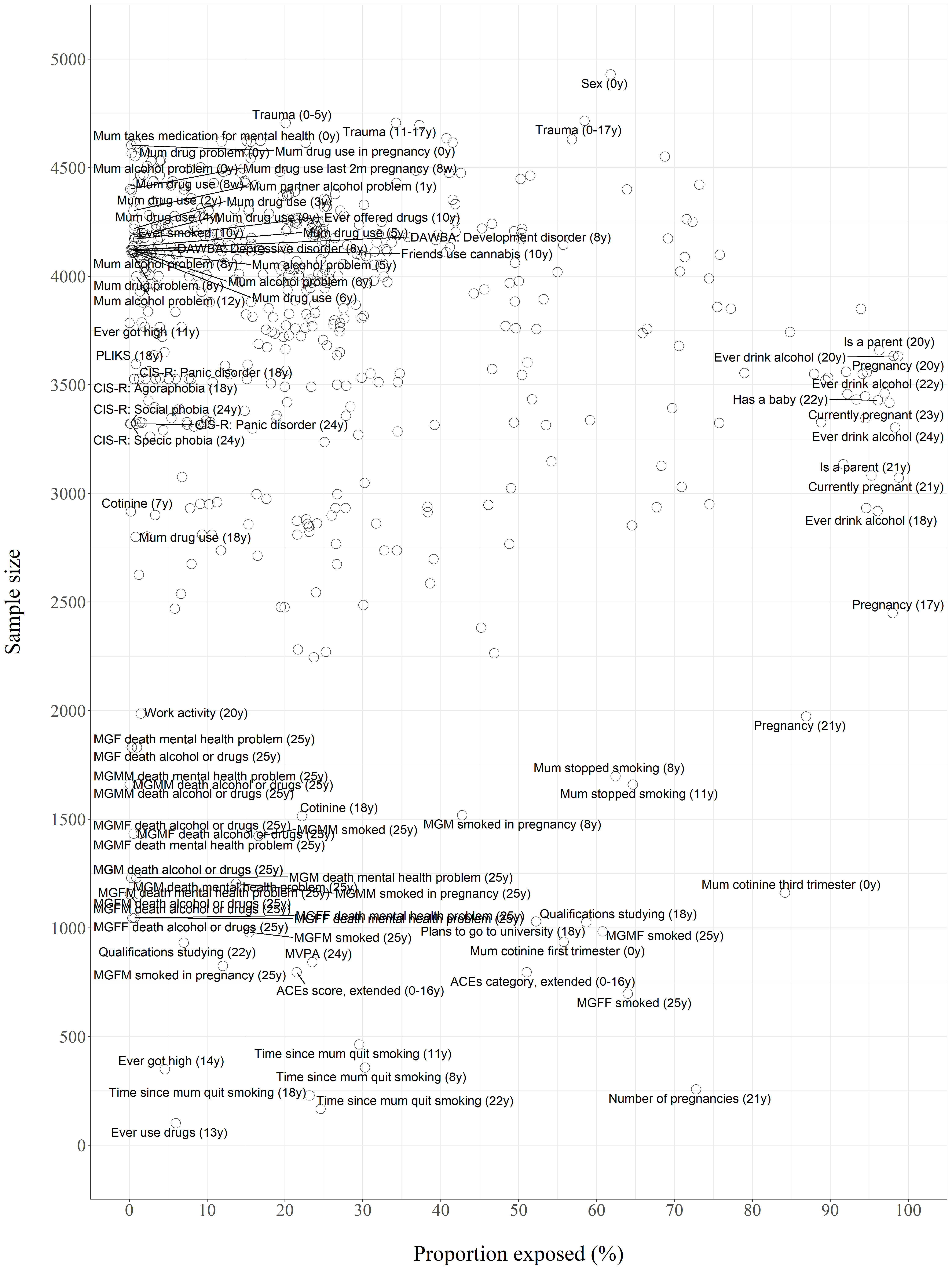

**Figure S6. Sample sizes for each risk factor by proportion exposed separated by comparison**

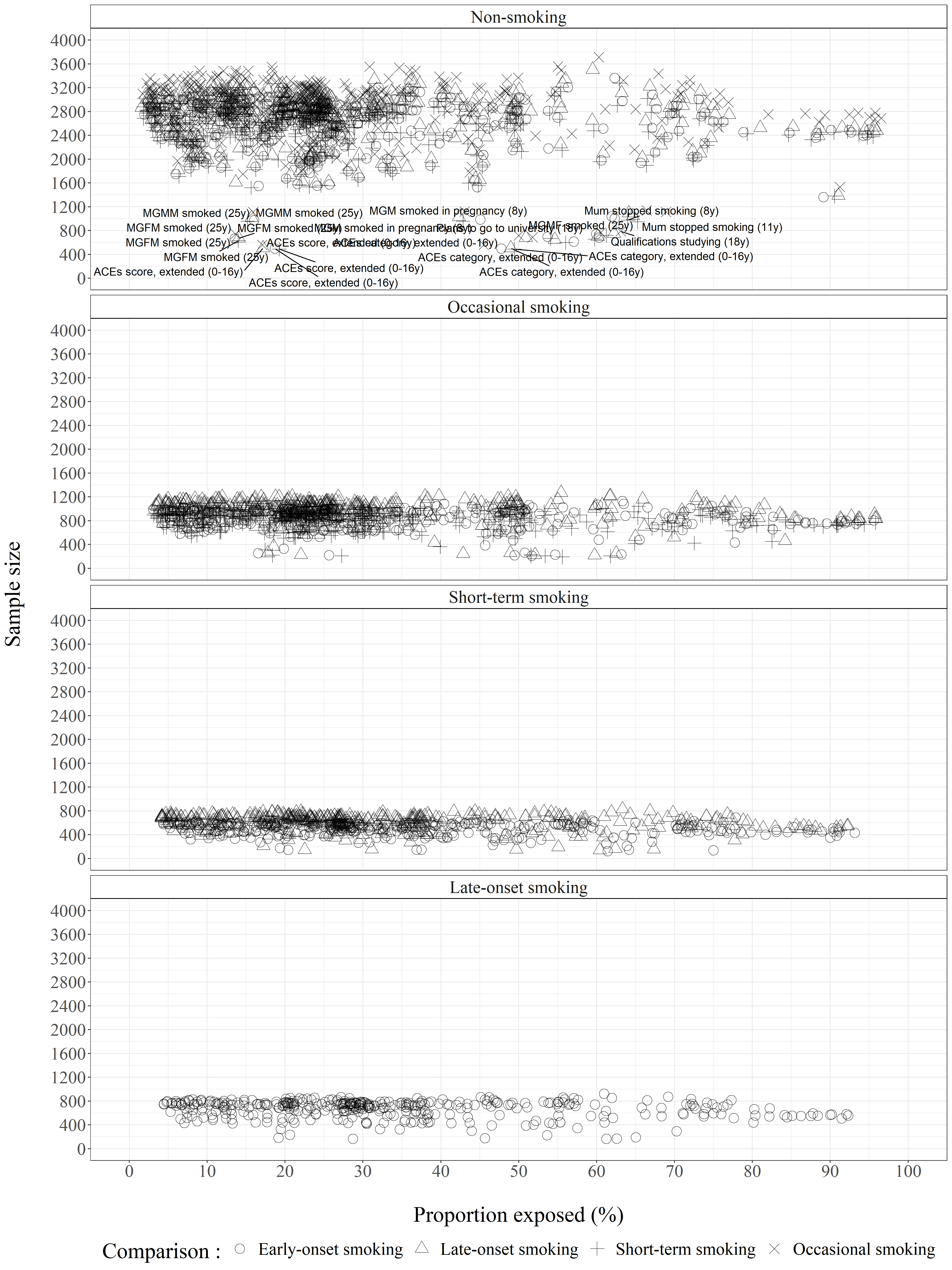

**Figure S7. Logged odds ratios by proportion exposed separated by comparison**

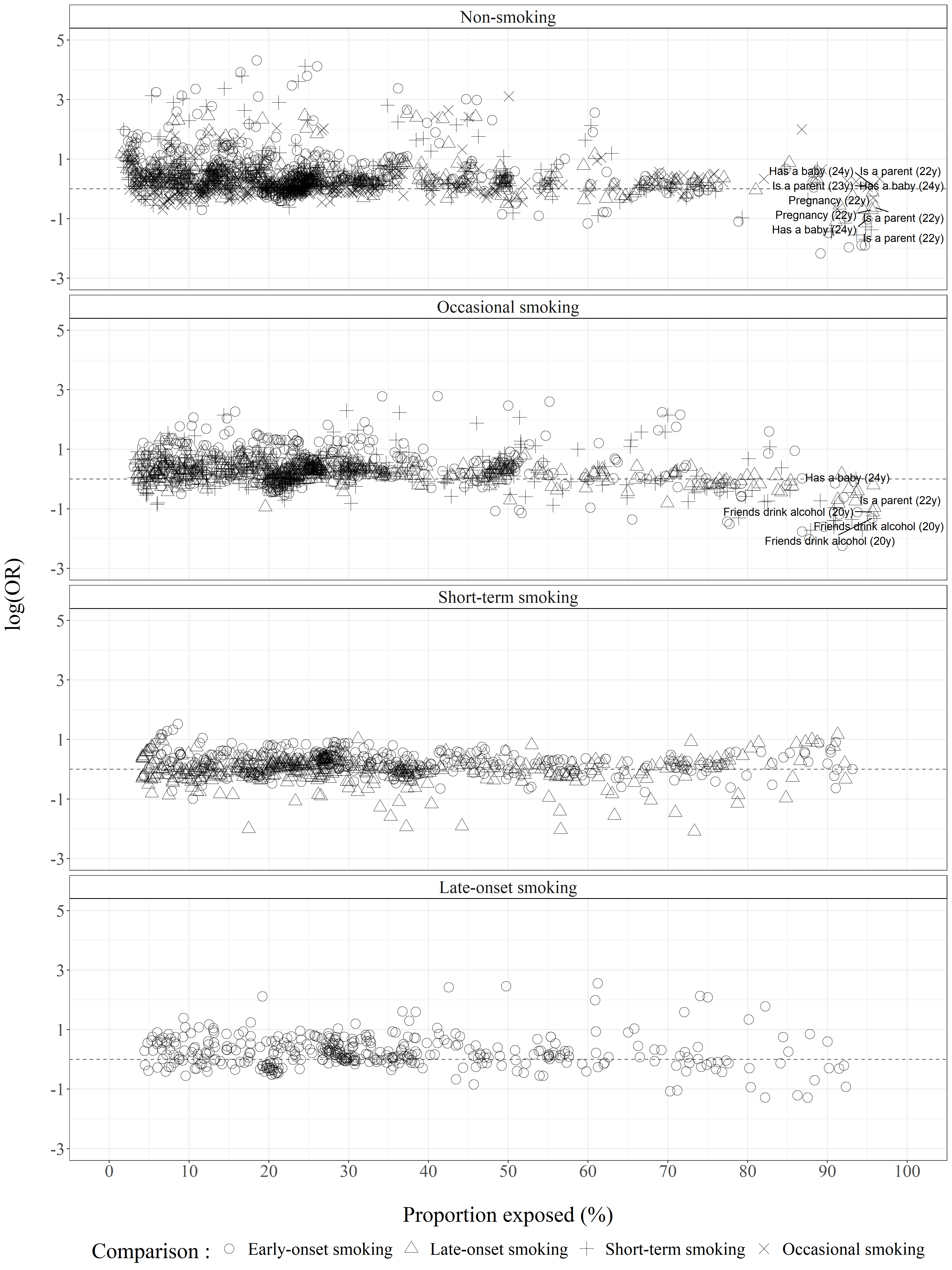

**Figure S8: Scatter plot showing log odds against population attributable fraction**

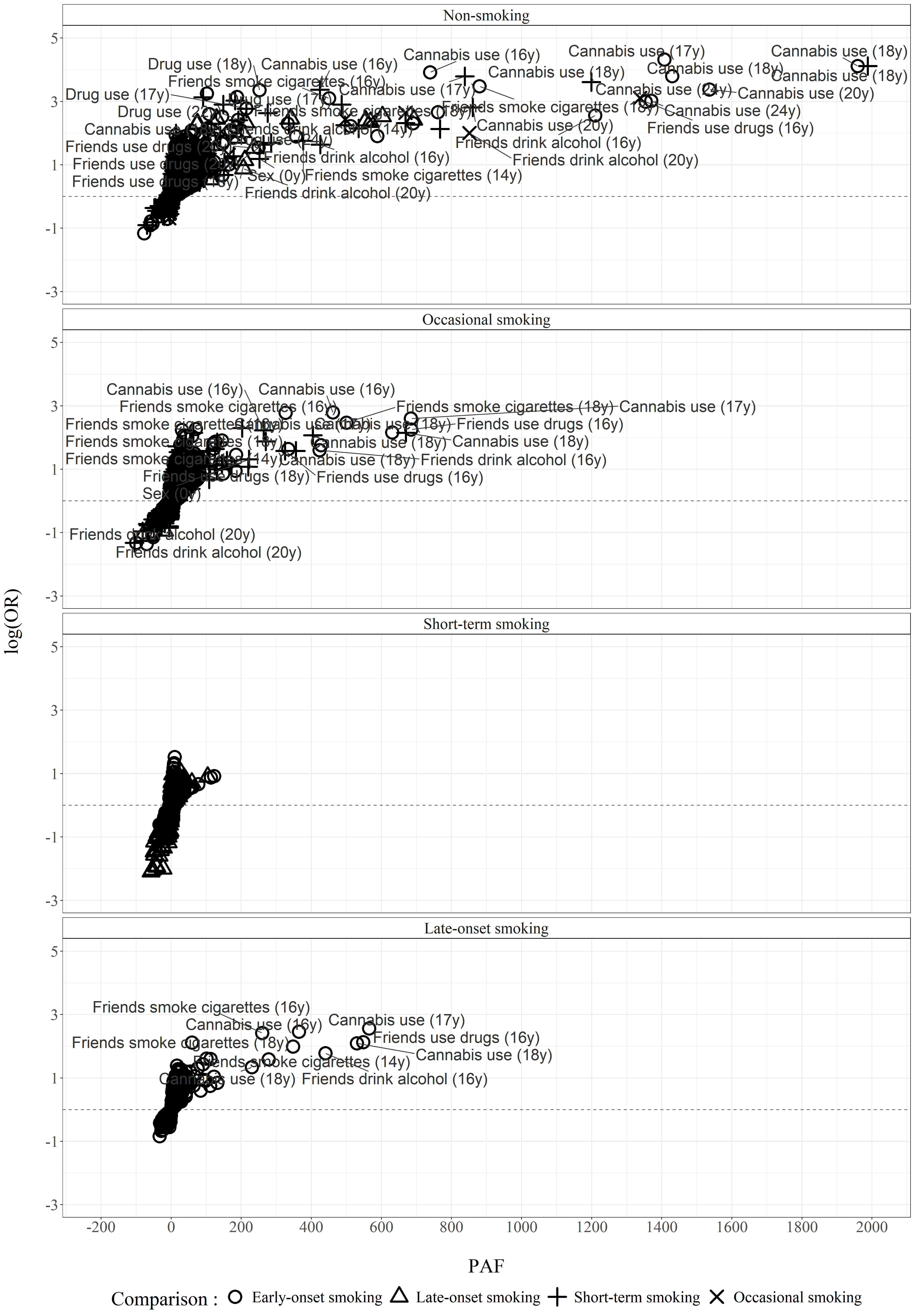

**Figure S9. Logged odds ratios by e-values separated by comparison**

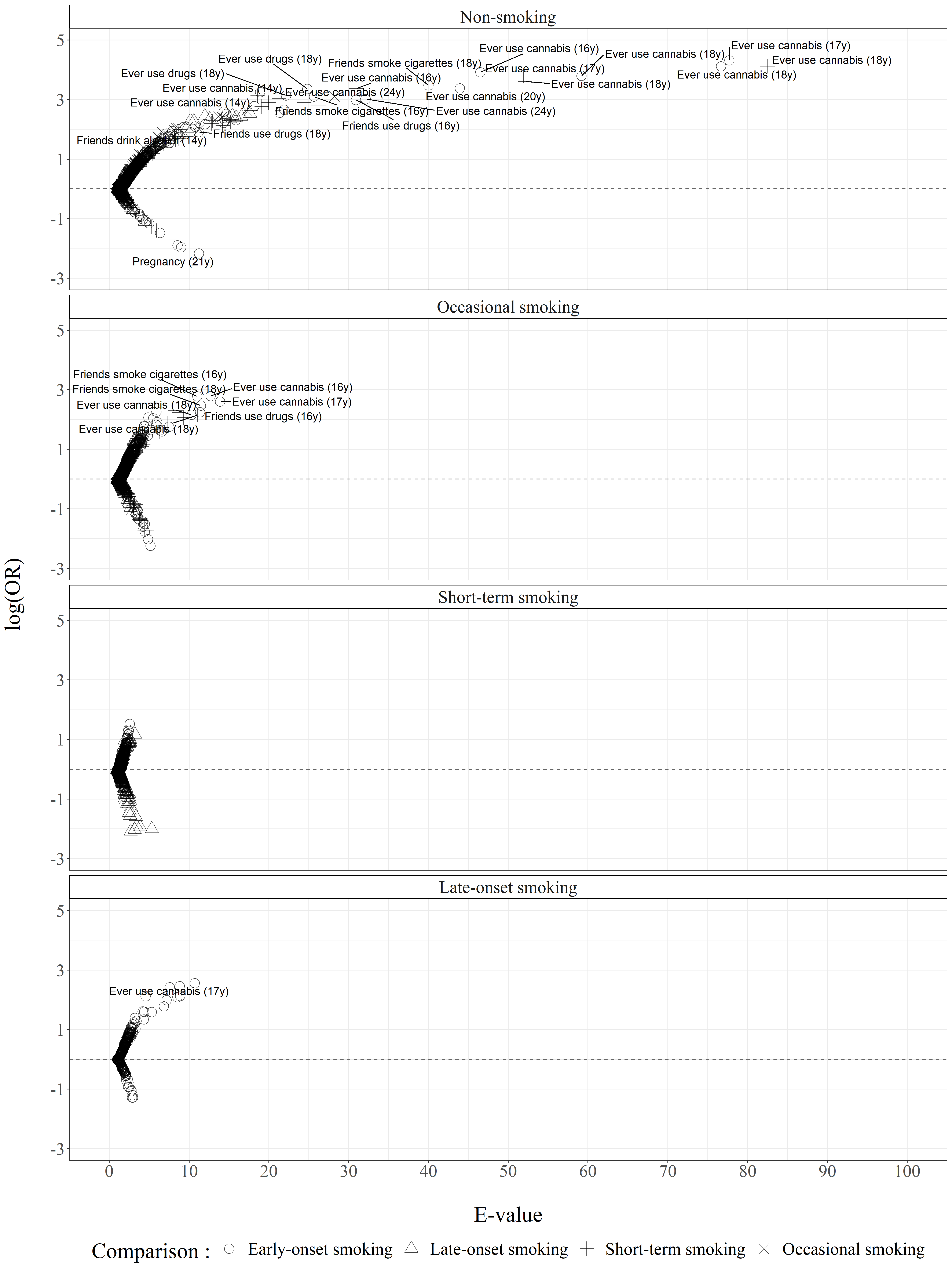

**Figures S10. Circle plots showing association between each risk factor measure and latent classes of smoking separated by exposure category**

**Figure S10.1. Non-smoking vs any smoking**

**Figure S10.1.1. Family substance use**

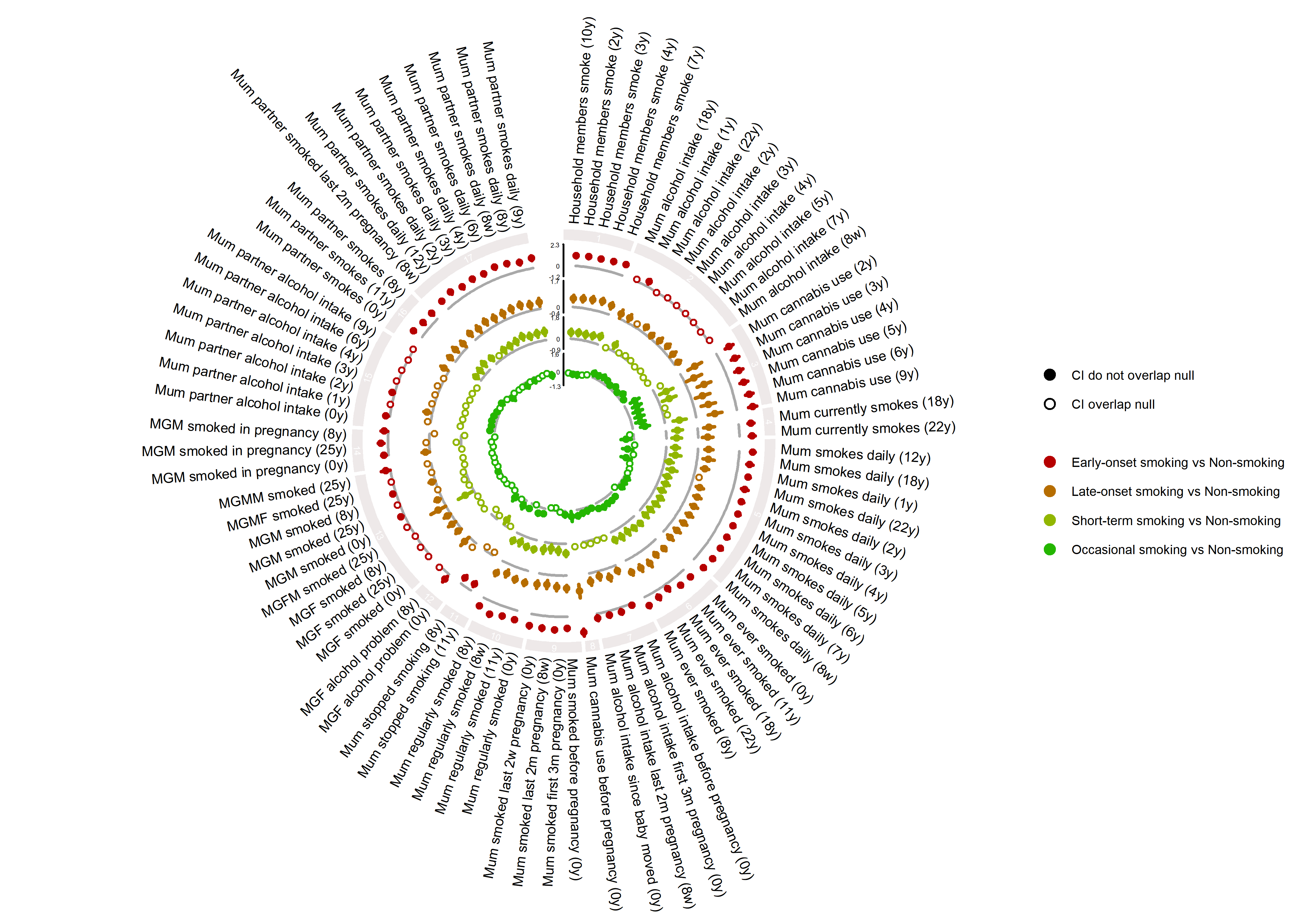

**Figure S10.1.2. Family sociodemographic factors**

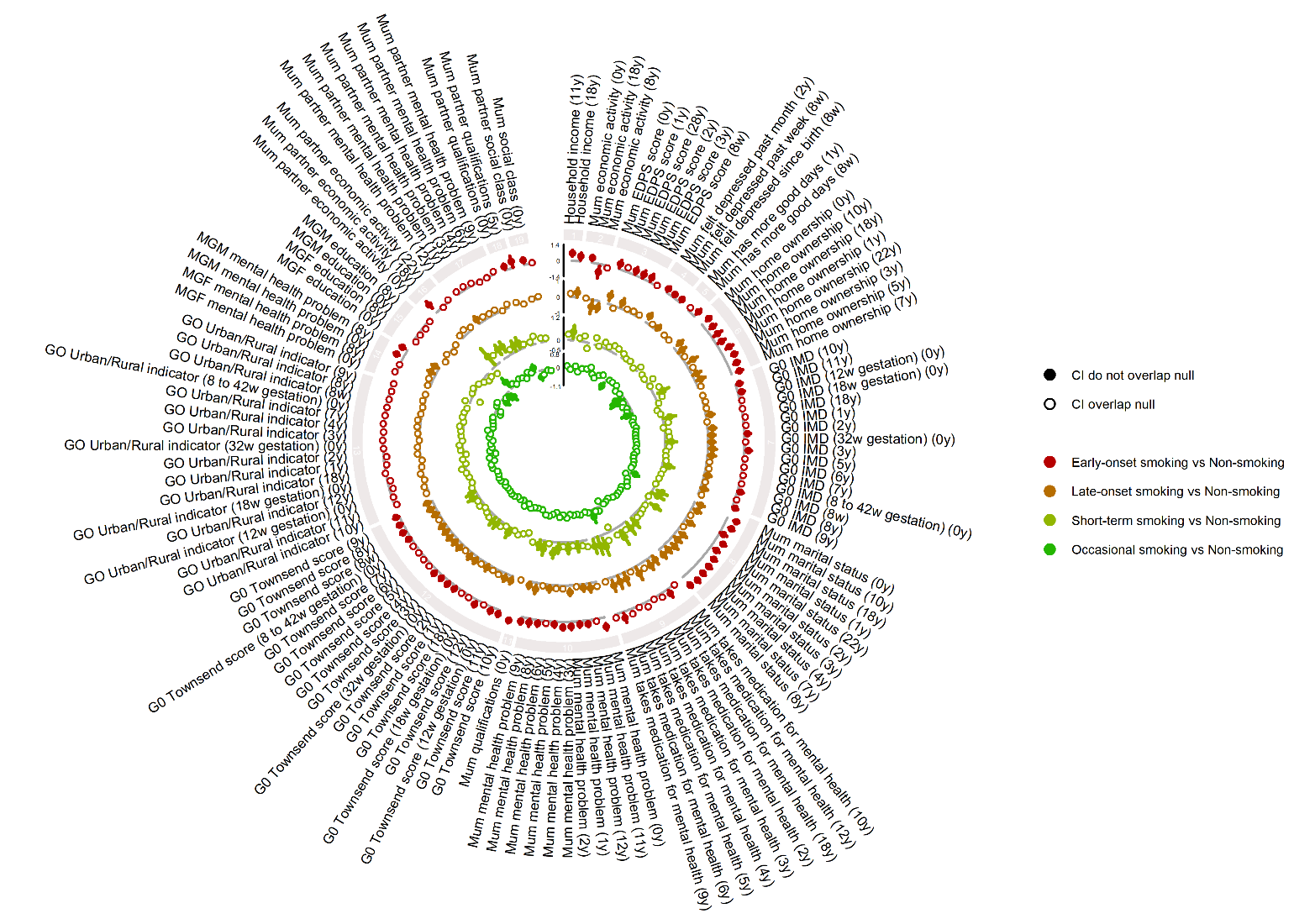

**Figure S10.1.3. Individual sociodemographic factors**

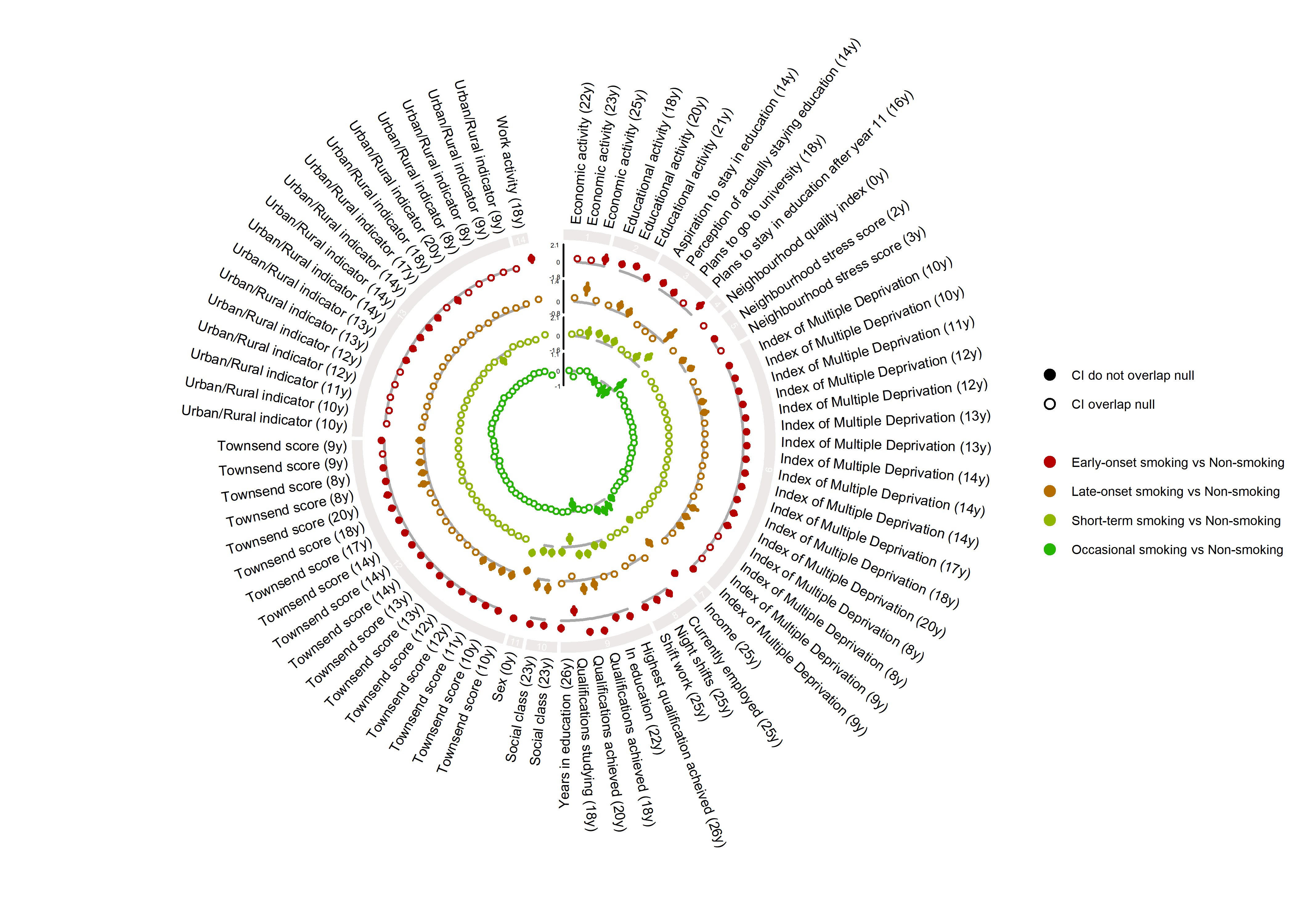

**Figure S10.1.4. Individual lifestyle and peer factors**

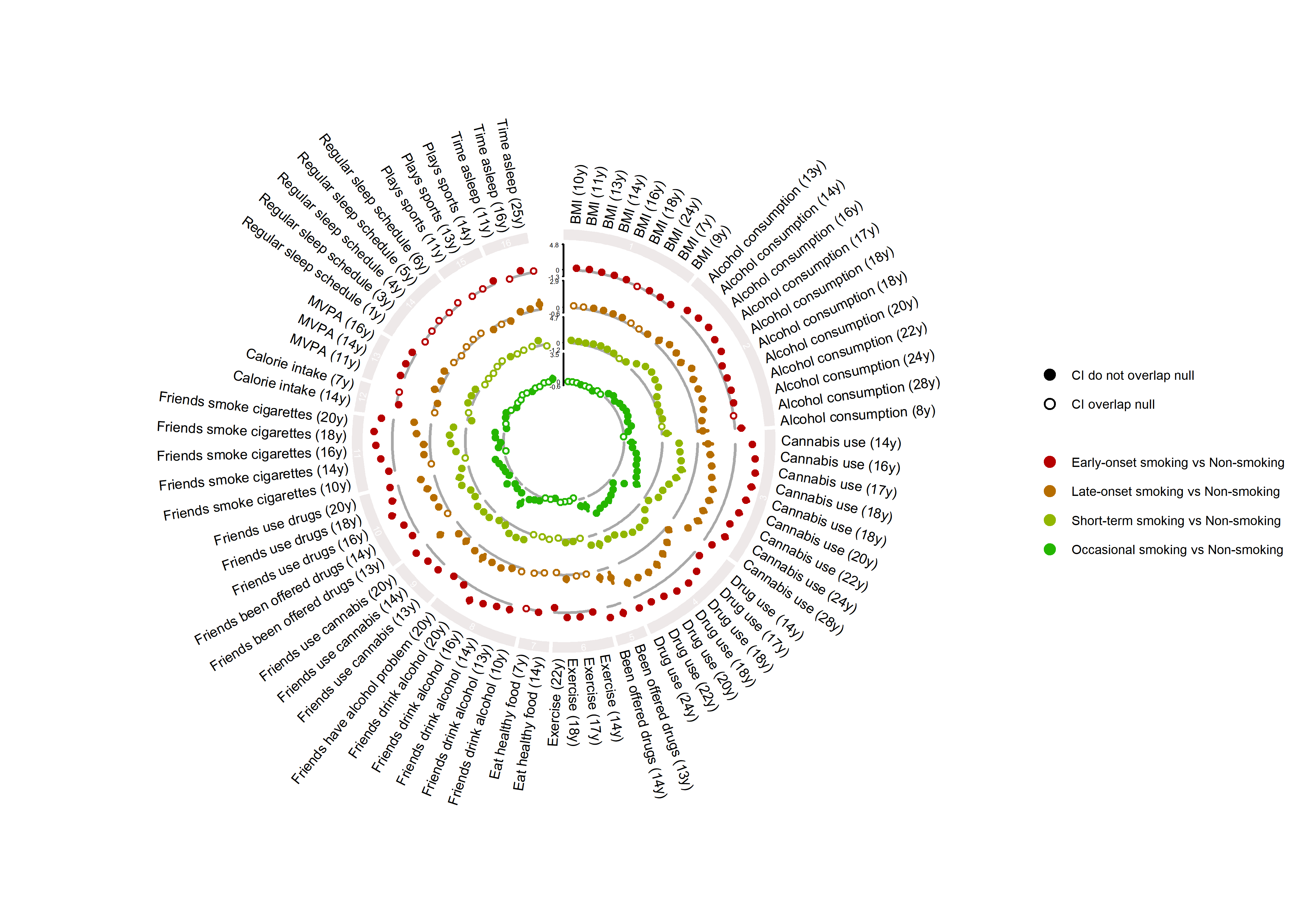

**Figure S10.1.5. Mental health and other factors**

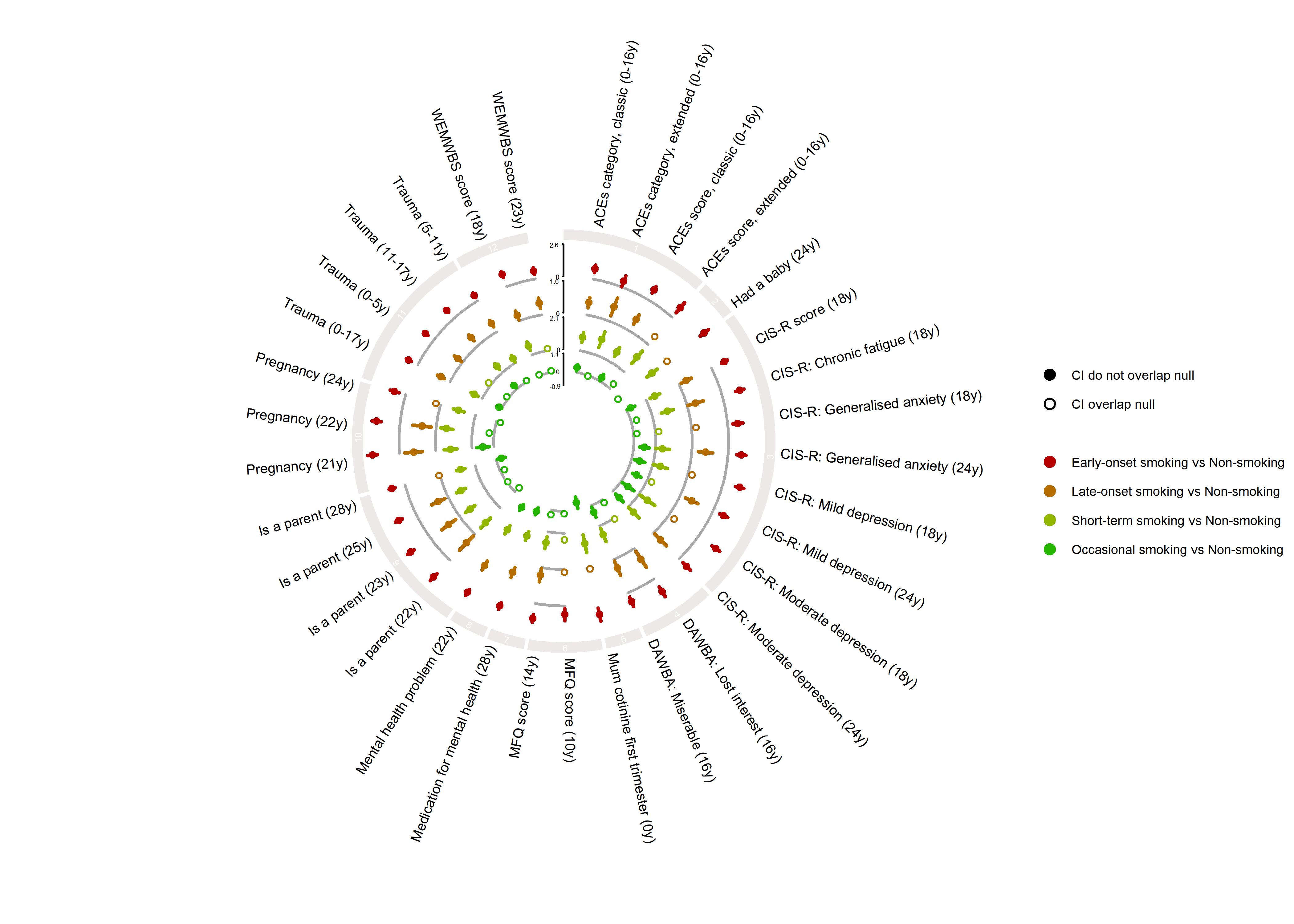

**Figure S10.2. Occasional smoking vs any regular smoking**

**Figure S10.2.1. Family substance use**

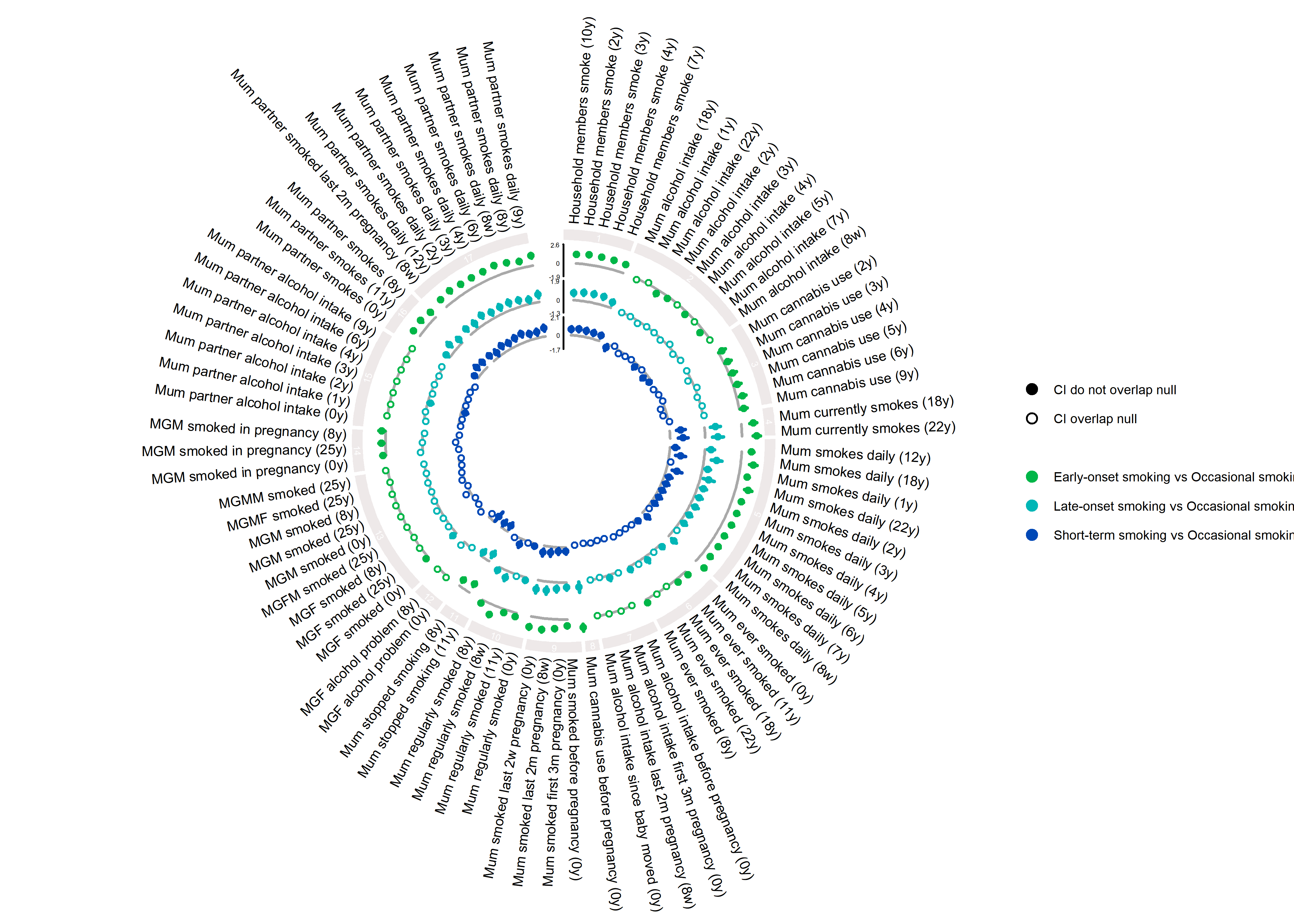

**Figure S10.2.2. Family sociodemographic factors**

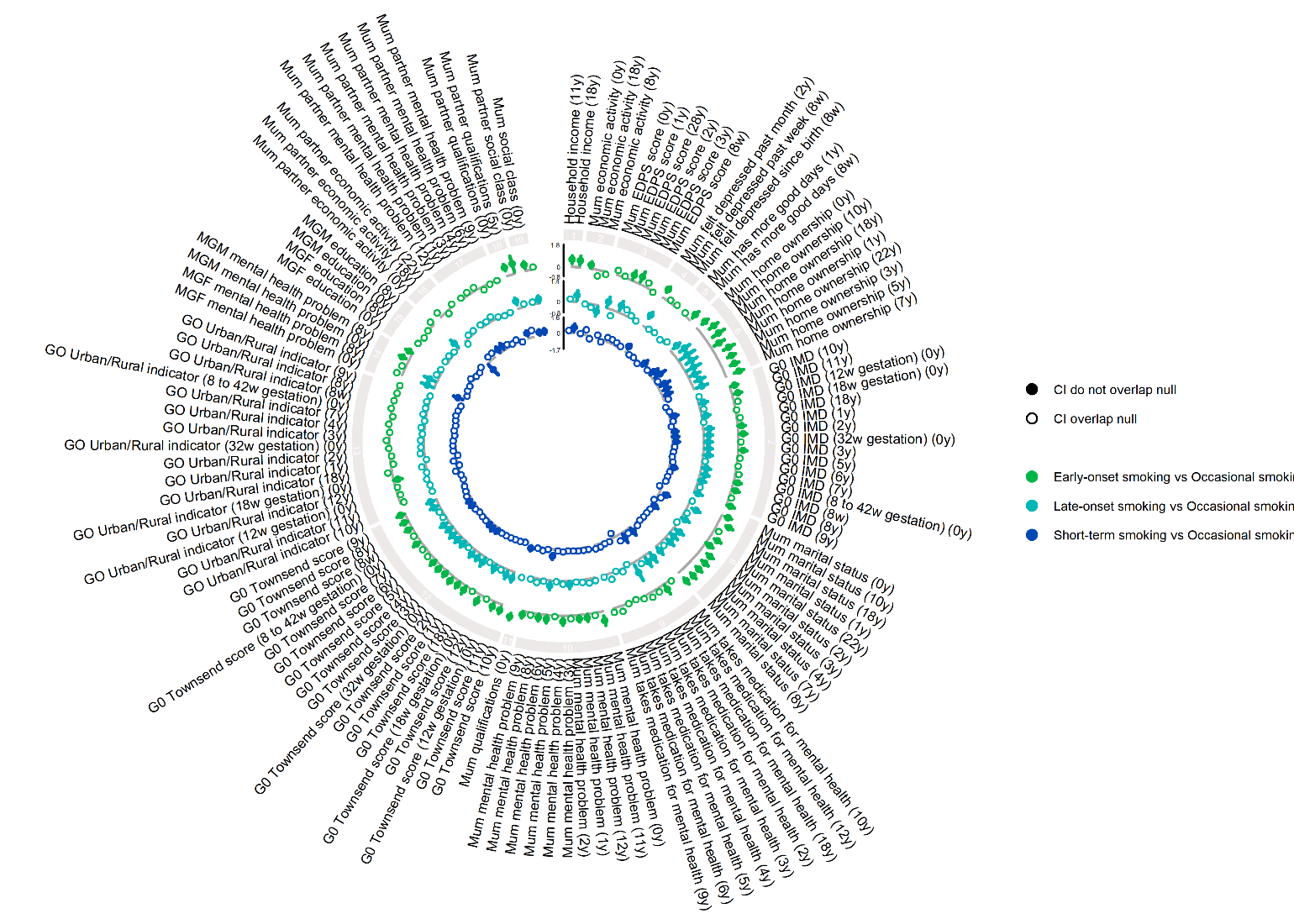

**Figure S10.2.3. Individual sociodemographic factors**

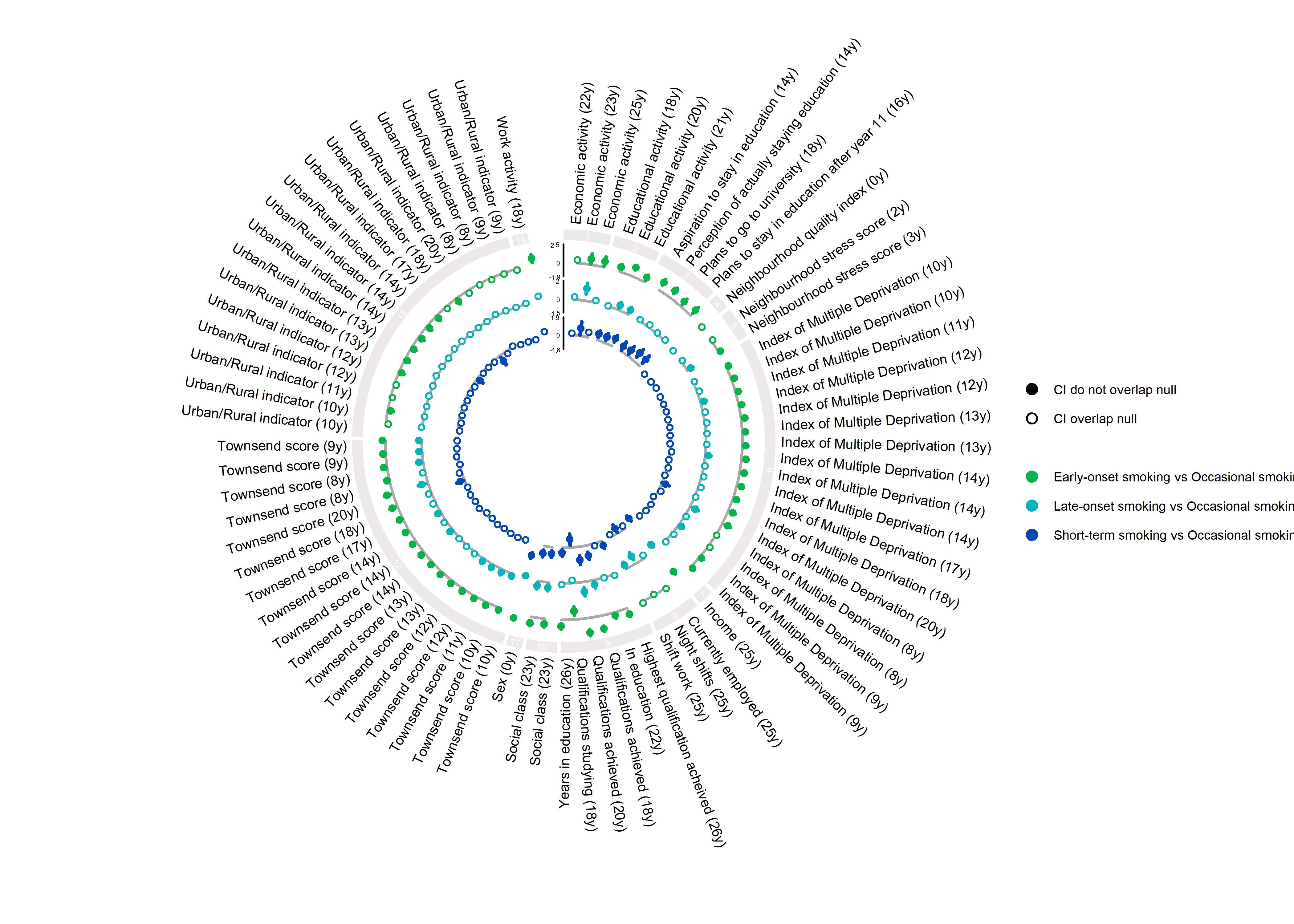

**Figure S10.2.4. Individual lifestyle and peer factors**

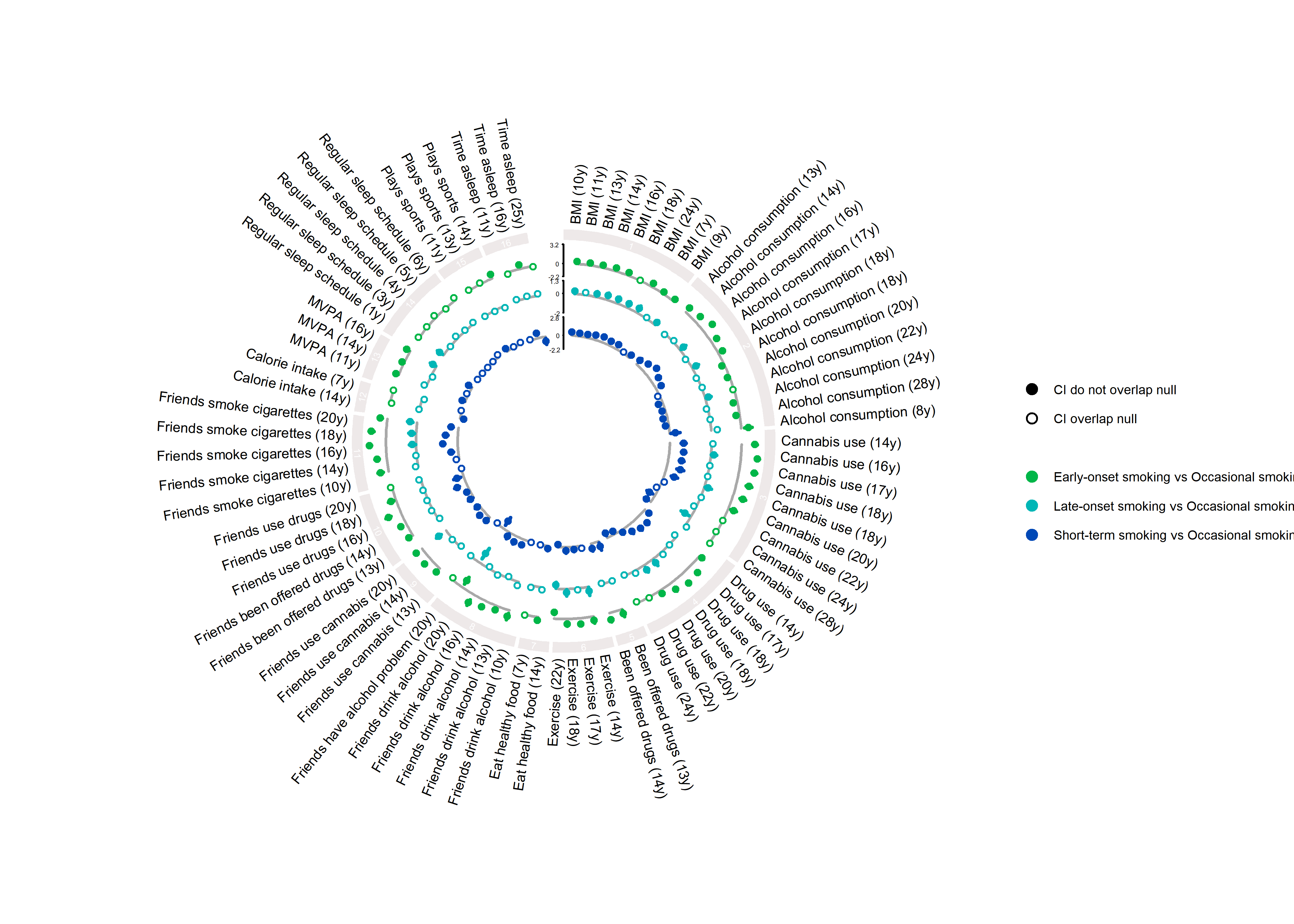

**Figure S10.2.5. Mental health and other factors**

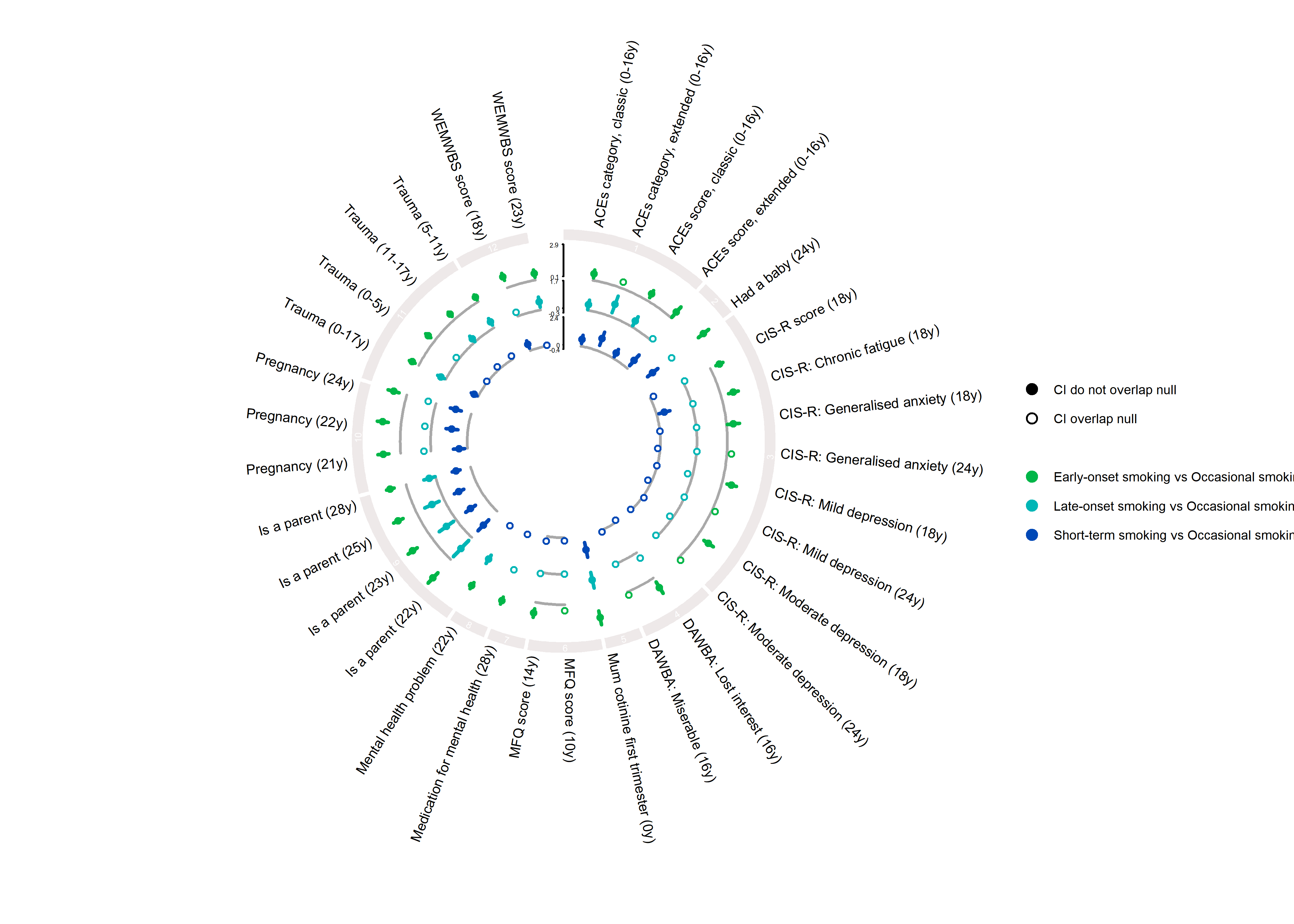

**Figure S10.3. Short-term smoking vs sustained smoking**

**Figure S10.3.1. Family substance use**

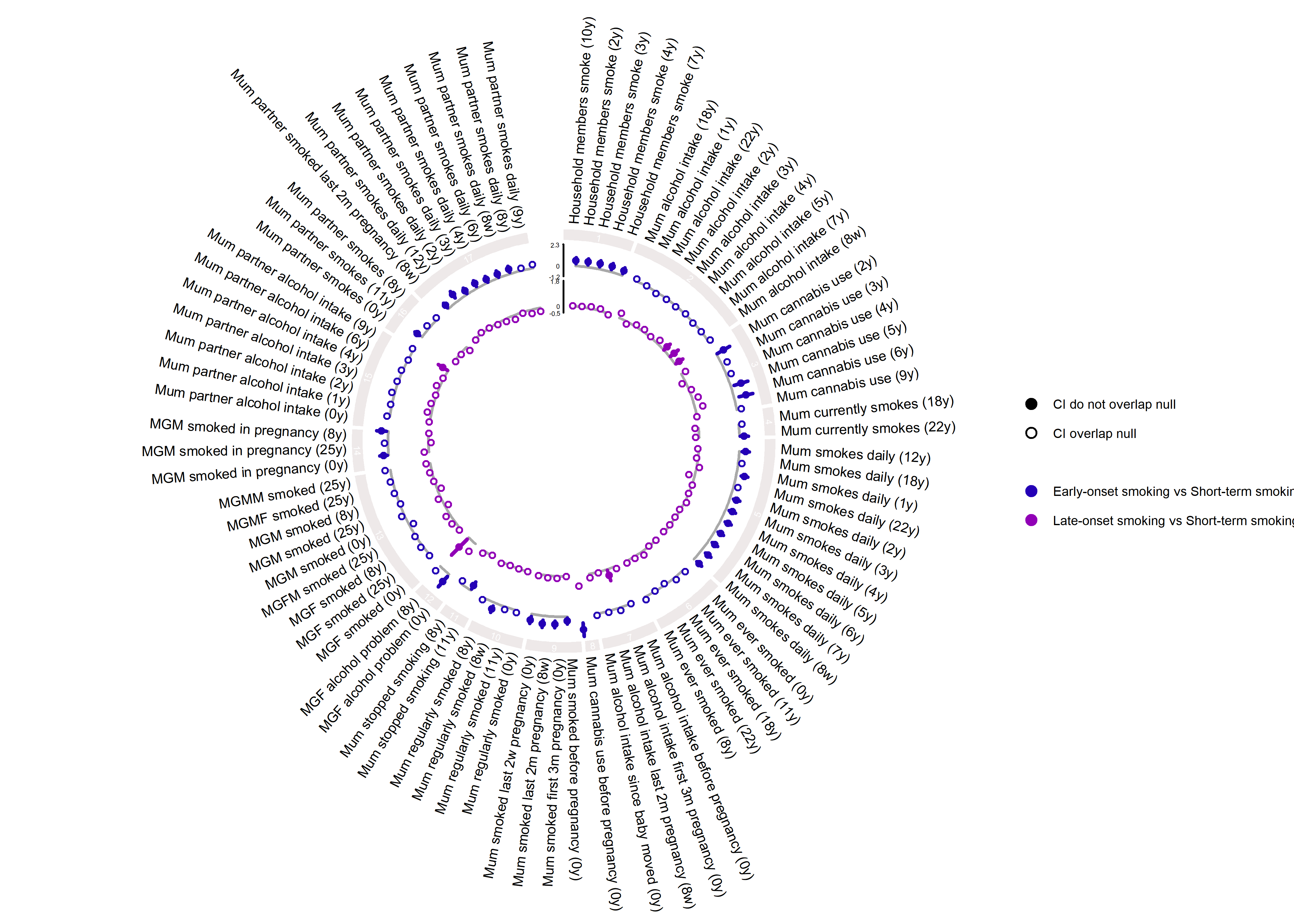

**Figure S10.3.2. Family sociodemographic factors**

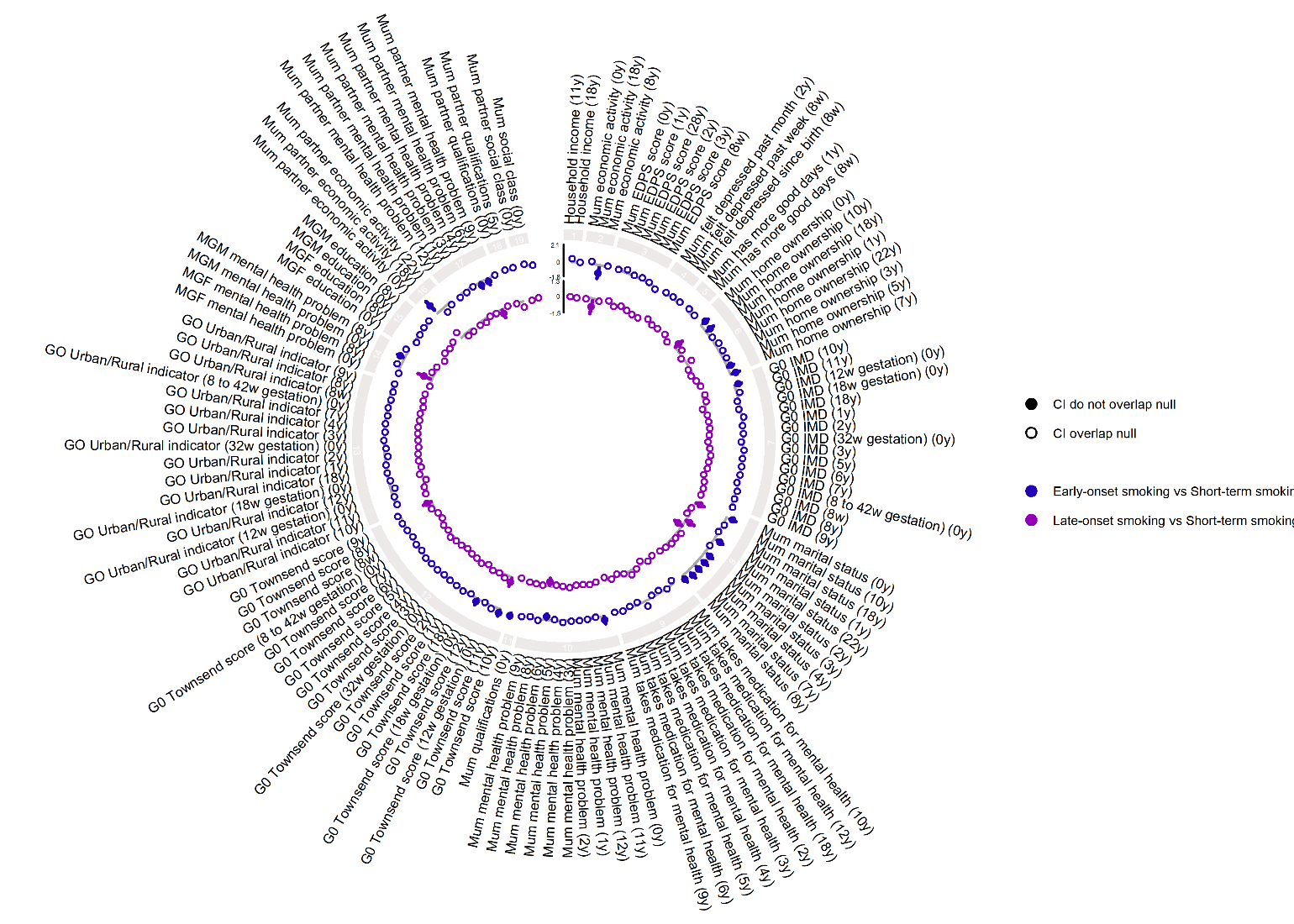

**Figure S10.3.3. Individual sociodemographic factors**

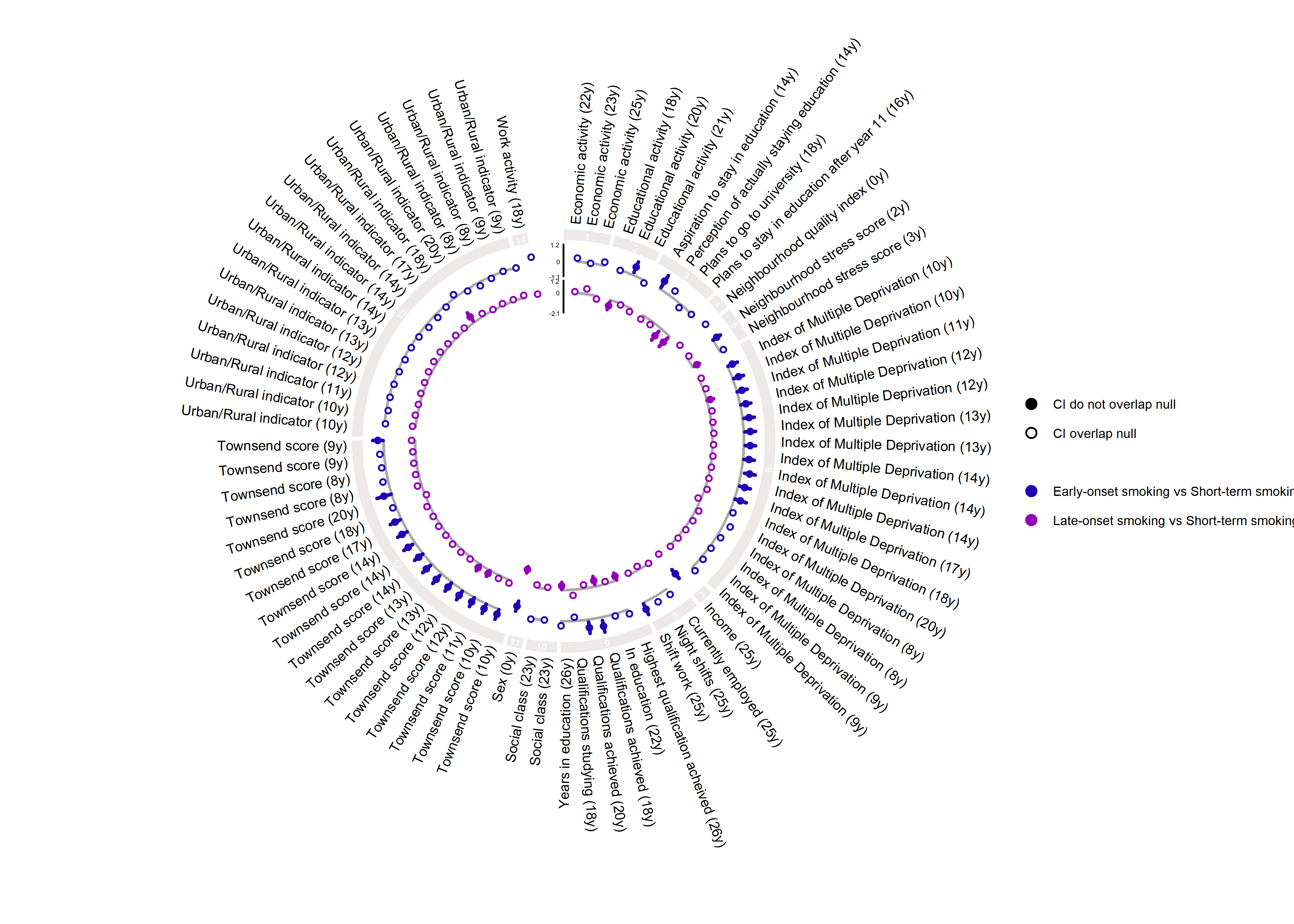

**Figure S10.3.4. Individual lifestyle and peer factors**

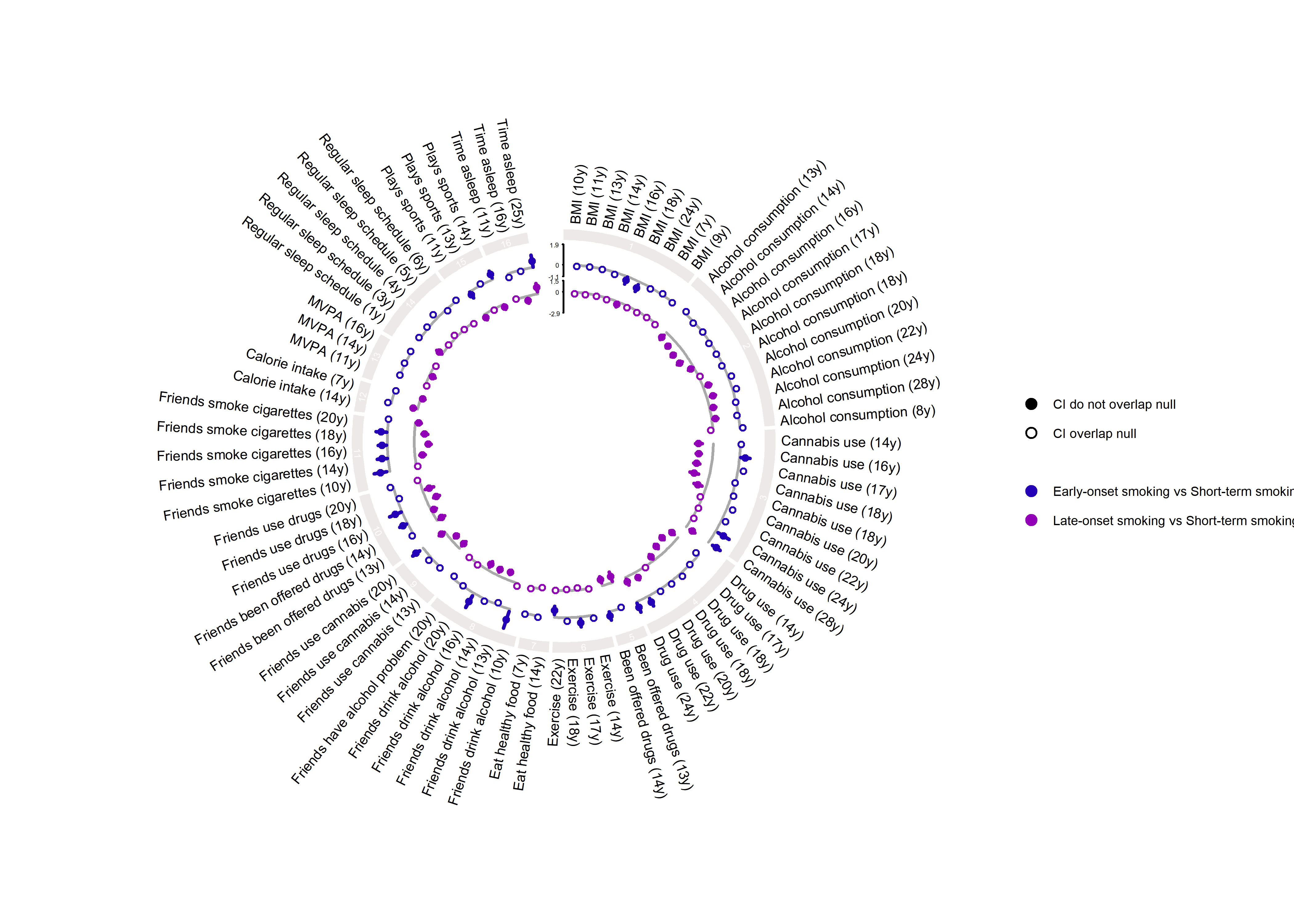

**Figure S10.3.5. Mental health and other factors**

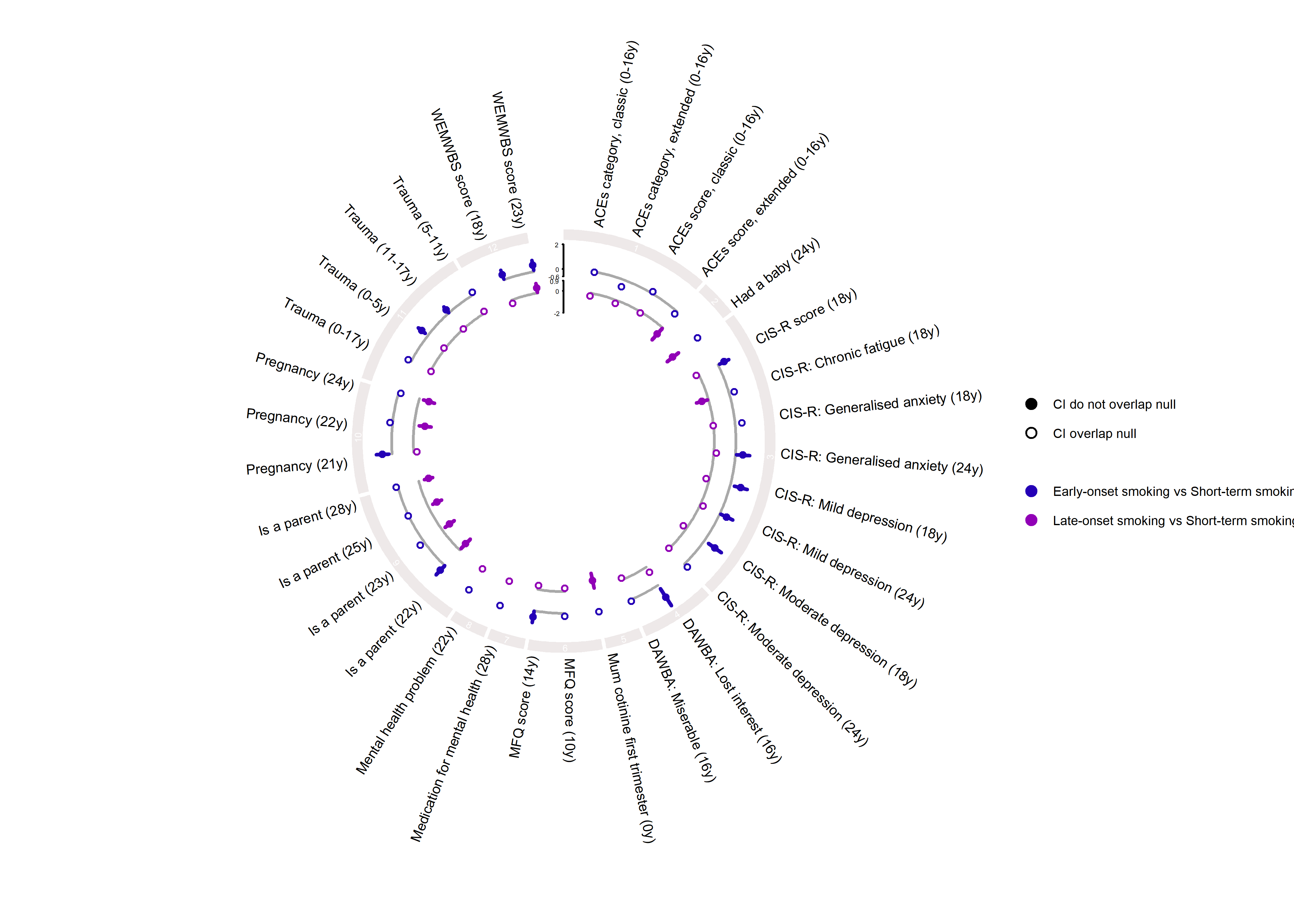

**Figure S10.4. Late-onset vs early-onset smoking**

**Figure S10.4.1. Family substance use**

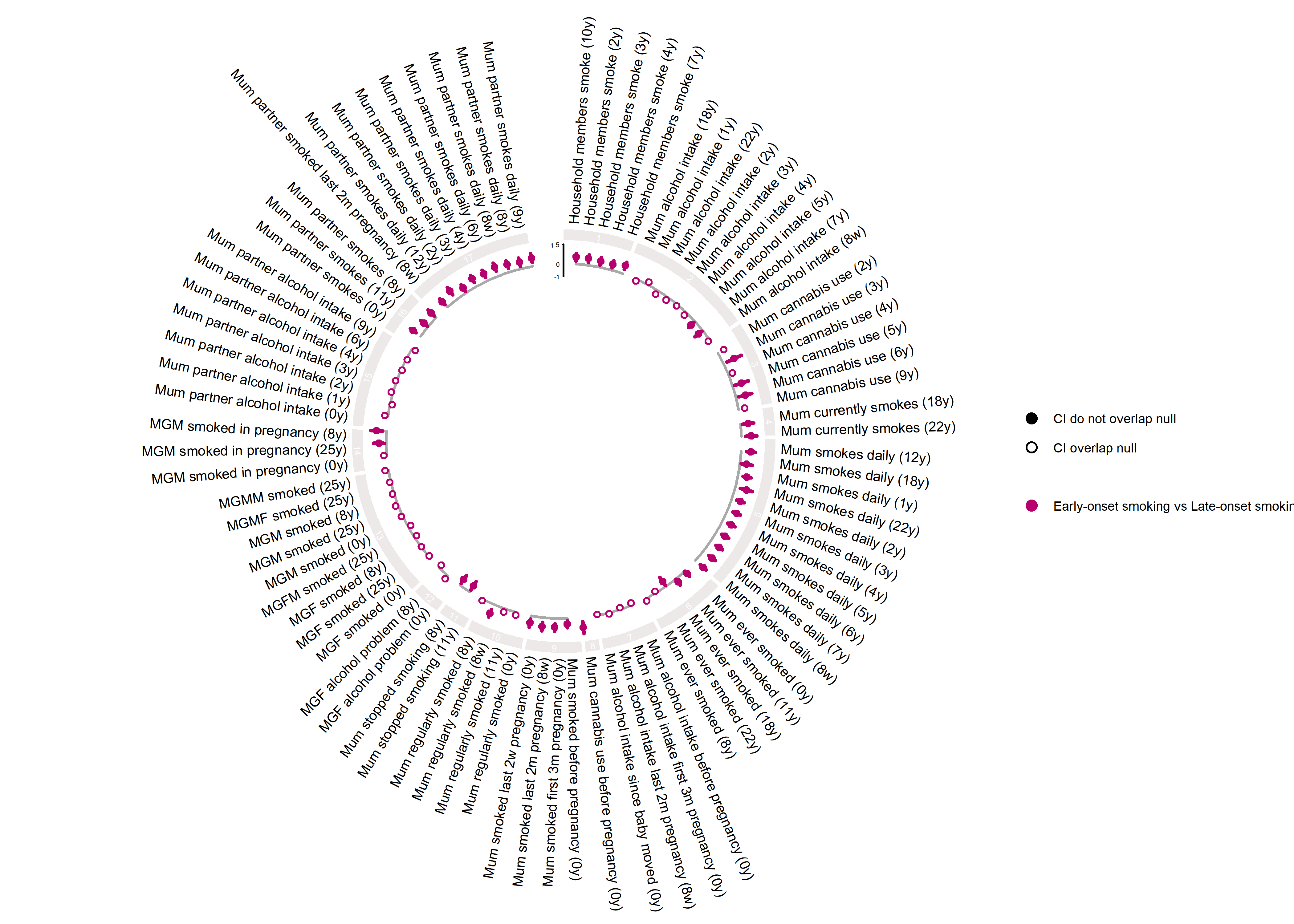

**Figure S10.4.2. Family sociodemographic factors**

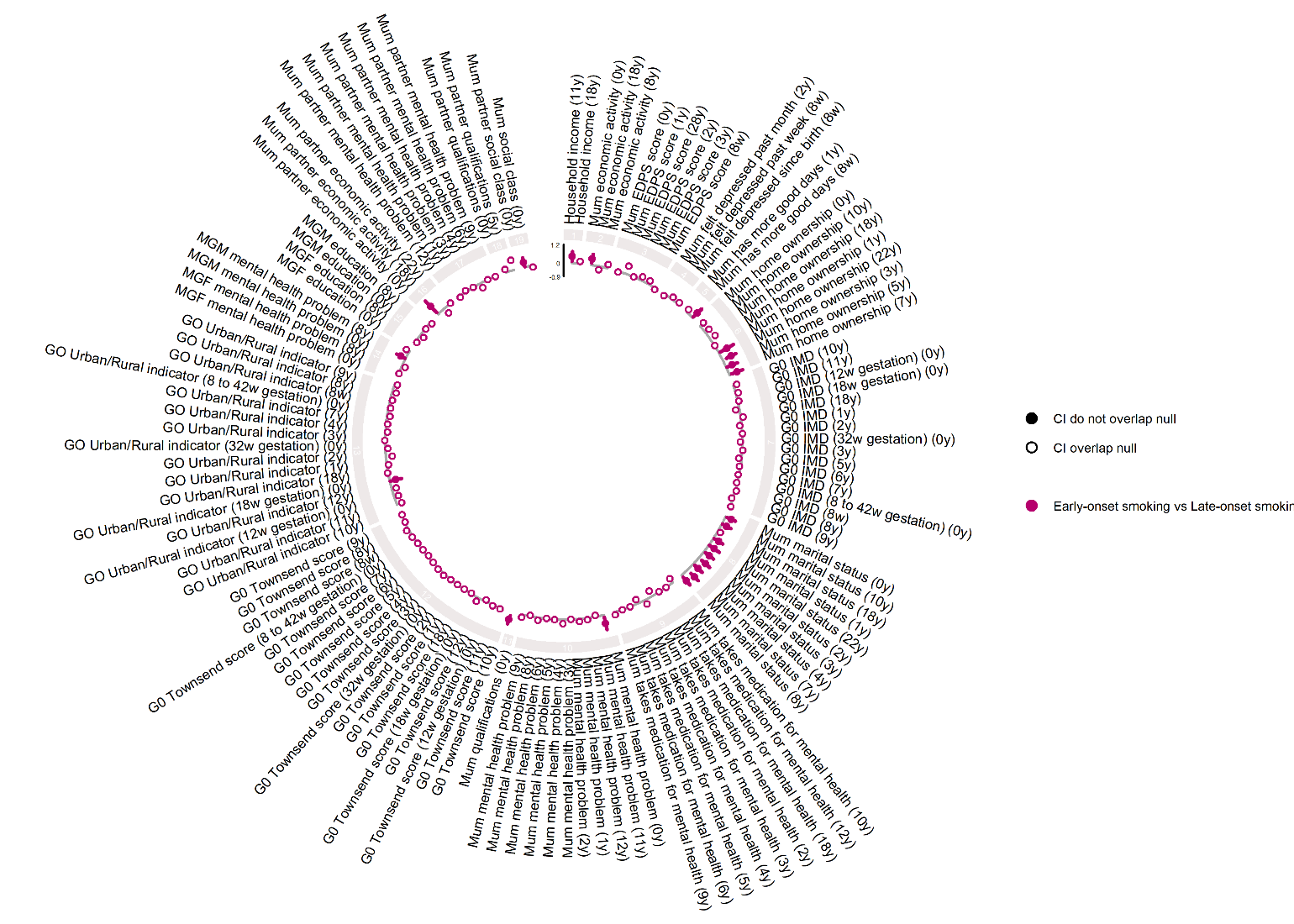

**Figure S10.4.3. Individual sociodemographic factors**

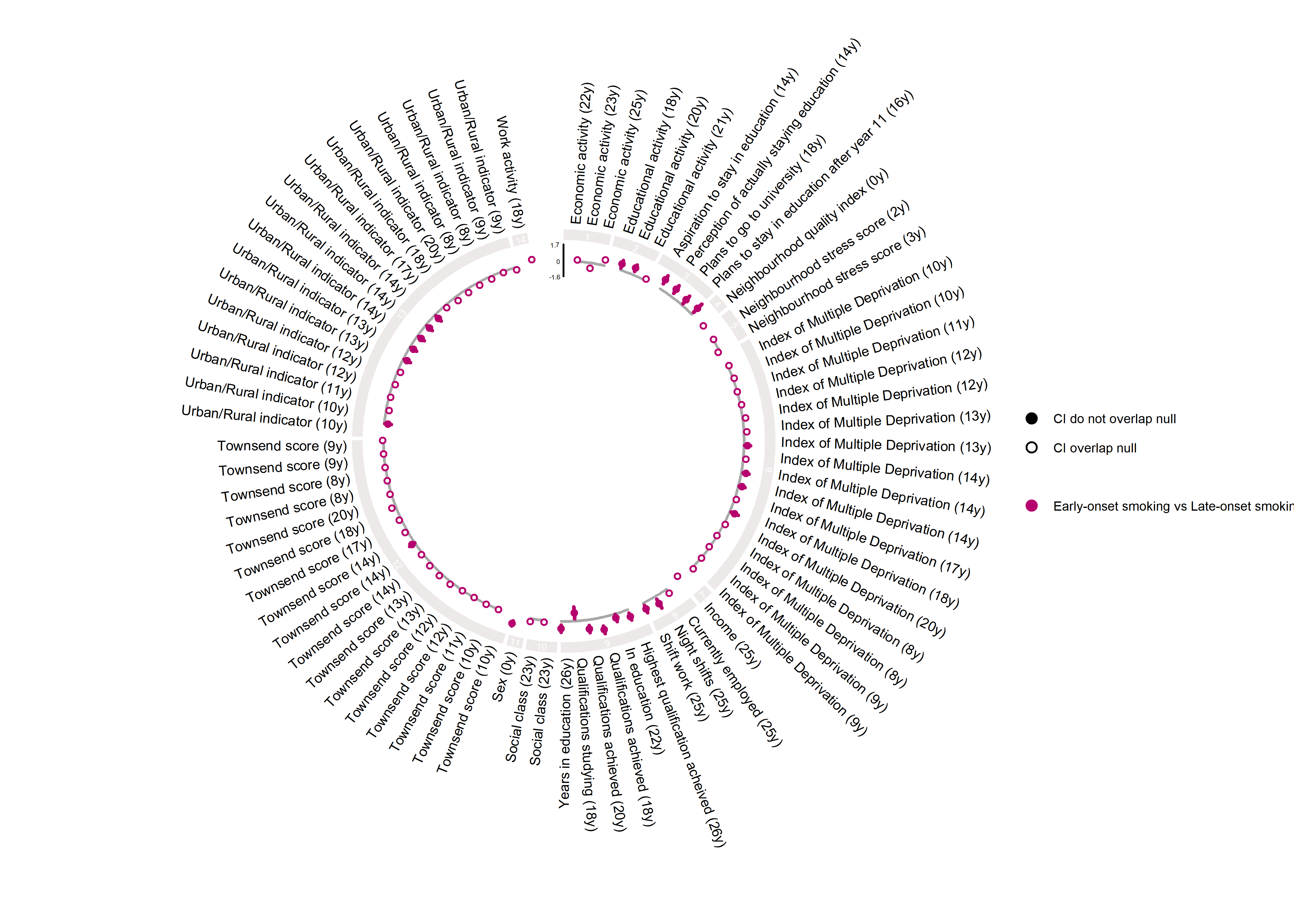

**Figure S10.4.4. Individual lifestyle and peer factors**

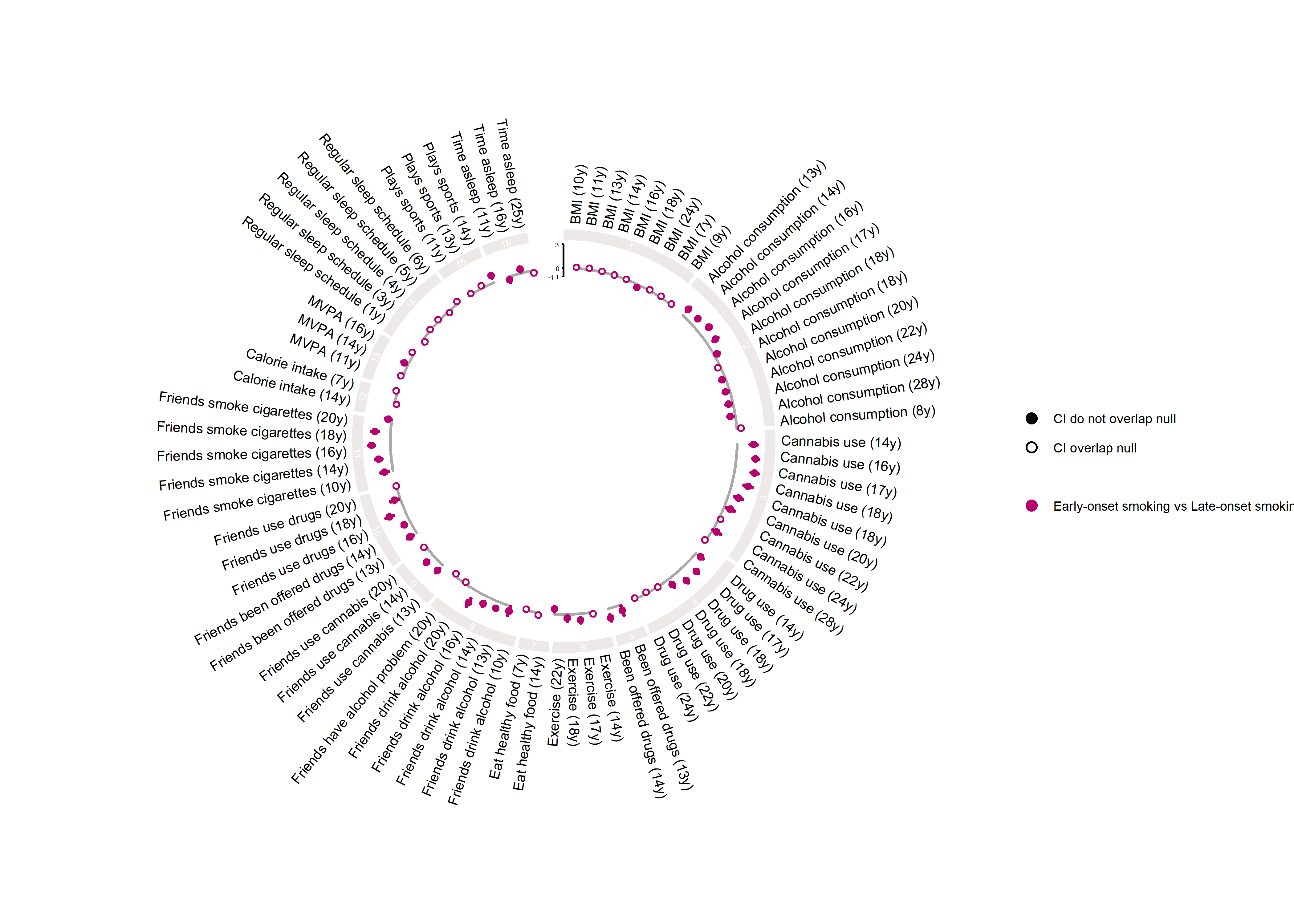

**Figure S10.4.5. Mental health and other factors**

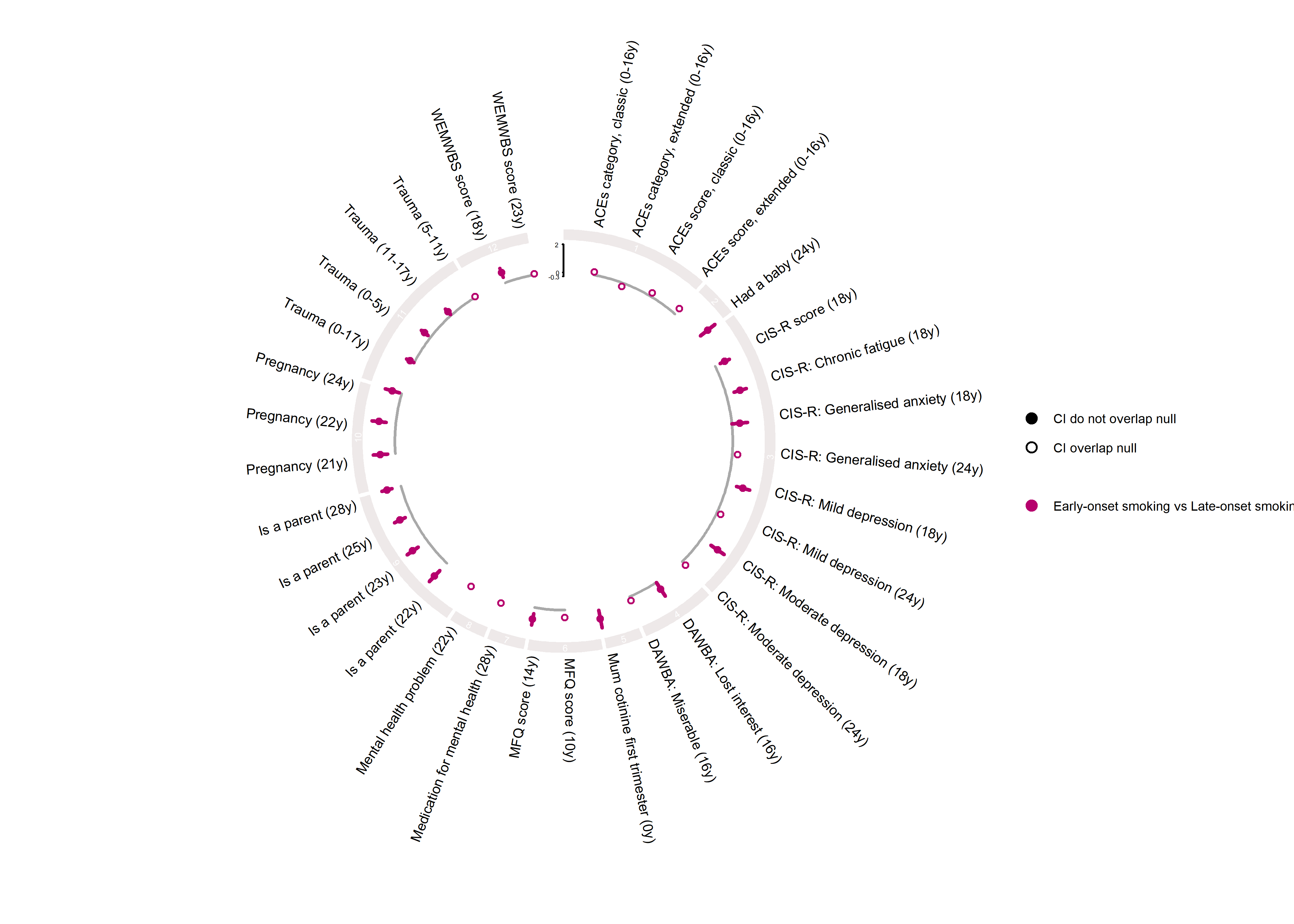
