## Supplementary Text for "Longitudinal patterns of smoking behaviours in adolescence and early adulthood and their association with modifiable and sociodemographic risk factors"

### 1. Smoking measures

Supplementary Table S1 shows the timepoints where each available smoking measure was collected in ALSPAC. Smoking variables collected at 12 regular intervals between the ages of 13 to 28 years (13, 14, 15, 16, 17, 18, 20, 21, 22, 23, 24, and 28 years) were used to construct a 4-category and 3-category smoking measure at each timepoint (Supplementary Table S2). This classifies participants as either (1) non-smoking, (2) occasional smoking, (3) weekly or (4) daily smoking, or combines weekly and daily smoking as (3) regular smoking. The variables used to generate smoking measures relate to ever smoking, current smoking and frequency of smoking. Timepoints are approximate for clarity. Average real ages and age ranges for each timepoint are shown in Supplementary Table S3.

A total of 15,645 participants took part in ALSPAC; 9,337 self-reported smoking in at least 1 timepoint and 4,937 participants self-reported smoking in at least 3 timepoints, with at least one measured in adolescence, one in young adulthood and one in their twenties. Supplementary Table S4 shows the number of participants who reported on their smoking behaviours at each timepoint or were missing. Most participants were non-smokers across all timepoints, and the lowest proportion of non-smokers is observed at the 21 year (CCU) timepoint. Supplementary Table S5 shows that the number of missing participants by the number of missing timepoints from 0 (no missing data) to 12 (all missing). 787 participants self-reported their smoking at all 12 included timepoints, most of whom were non-smokers. This shows that earlier timepoints generally show less missingness than later timepoints, with an exception at 24 years where questionnaire data was supplemented with clinic data. The timepoint with the greatest amount of missing smoking data is at 22 years when including all participants and at 19 years when limiting to participants with at least 3 non-missing timepoints.

### 2. Exposure measures

Factors of interest include familial and peer influences, sociodemographic factors, lifestyle factors, mental health, deprivation and more. Supplementary Tables S6-S7 show the time points where each risk factor measure was collected. Sociodemographic factors investigated include the participant’s sex assigned at birth, ethnicity of the participants and parents, familial education, parental SEP, education, employment, and neighbourhood deprivation. The modifiable risk factors of interest include familial smoking and substance use, familial mental health, peer substance use, other substance use, mental health and wellbeing, body mass index (BMI), diet, physical activity, and sleep. Other risk factors of interest include adverse childhood experiences (ACEs), trauma, pregnancy or parenthood, and cotinine. Cotinine is a metabolite of nicotine and can be used to biochemically assess smoking. Measures of adverse childhood experiences (ACEs) and trauma were previously derived longitudinally (Houtepen et al., 2018; Croft et al., 2019). Measures of neighbourhood deprivation were derived from postcodes (Noble et al., 2006; Morris et al., 2018).

Modifiable risk factors present potential intervention targets and sociodemographic factors could be used to target public health policy and prevention programmes in relation to smoking. Modifiable risk factors may also in turn be shaped by sociodemographic factors.

Risk factor measures collected at timepoints up to 13 years, preceding the derived smoking patterns, represent baseline risk factors while measures collected after 13 years represent concurrent risk factors. The direction of association for concurrent measures is less clear than for preceding measures as some concurrent risk factors occur before or at the same time as many of the smoking measures.

### 3. Latent class analyses

Latent class analysis was used to characterise changes in smoking behaviour from early adolescence through to adulthood. This allowed for the identification and classification of separate subgroups of participants with similar patterns of behaviour. The resulting latent classes are dependent not only on the data used but also differences in timing, spacing, number of measurements available, categorisation of smoking used, and various model selection and fit criteria. Given that 12 timepoints are used here, when using a 4-category smoking measure a total of 16,777,216 response patterns are possible while using a 3-category a total of 531,441 response patterns are possible.

Mplus 8 was used to fit latent class models and multiple random starting values were utilized to avoid convergence on local maxima when fitting the model to calculate the maximum likelihood. Individuals with missing data were included in the analysis using full information maximum likelihood which assumes missing at random. This means missingness may be related to the smoking data used but not to the missing data itself.

Longitudinal latent class analysis (LLCA) was used to model the associations between repeated measures of smoking. This uses a multinomial logit model that fits thresholds to model patterns of change between timepoints and does not assume proportional odds. This aims to elucidate latent classes within which the repeated measures of smoking are no longer associated making them conditionally independent. Models were fit with an increasing number of classes until the model with the best fit to the data was determined. The optimal number of latent classes that adequately explained the relationship between the repeated measures was selected using sample size–adjusted Bayesian information criterion, the proportion of individuals in each class, entropy, convergence, and interpretability (Asparouhov and Muthen, 2013).

Information criterions are a relative measure used to assess the fit of a model while considering parsimony. The number of classes showing the lowest value of the information criterion is preferred. Ideally the information criterion will decrease in value as the number of classes increase until the optimal number of classes can be identified. However, in the instance that the information criterion continues to decrease the optimal number of classes can instead be decided by which model contributes a substantial decrease in the information criterion and the addition of more classes leads to only slight reductions. Entropy is a measure of how certain an individual’s class assignment is based on the posterior probability of class membership where less uncertainty signifies a better fit to the data.

**LLCA using 4-category smoking data**

Supplementary Table S8 shows fit statistics for the LLCA models fitted with 2 to 7 classes. When using all participants with at least one self-reported measure of smoking and 4-category smoking measures, all models show an entropy below 0.8 which indicates poor class separation. When using participants with at least 3 non-missing timepoints all models show entropy greater than 0.8 and better class separation. Using 4-category smoking measures, the information criterion is lowest for the six-class model. These latent classes are visualised using stacked bar graphs in Supplementary Figure S2 which shows the predicted probability of smoking by age for each of the six classes. These six classes are described as (1) early-onset smoking, (2) late-onset smoking, (3) non-daily smoking, (4) light smoking, (5) short-term smoking, and (6) non- smoking. Non- smoking was the largest class (62). Light smoking was the second largest class (15%) where the probability of smoking weekly or daily was near zero across all timepoints. With light smoking the probability of occasional smoking gradually increased in adolescence and then decreases in their twenties however across all timepoints most of the light smokers were likely to not smoke. The late-onset (7%) and early-onset (7%) classes were of similar size, and both consisted largely of daily smokers. Late-onset smoking initially shows a low probability of smoking and then the probability of daily smoking increased over time whereas in the early-onset smoking class participants were likely to smoke daily at an earlier age. Non-daily smoking was the fifth largest class (5.2%) and participants in this class are likely to smoke occasionally or weekly but not daily and appear to initiate smoking at a similar time as the late-onset smoking class. Short-term smoking was the smallest class (3.8%) and here smoking appears to increase gradually up to 17/18 years and then decreases thereafter, suggesting smoking cessation in early adulthood. In the five-class solution using 4-category smoking a latent class related to short-term smoking is not seen.

**LLCA using 3-category smoking data**

Using 3-category smoking measures required fewer parameters and as such the information criterions are lower for these models compared to similar models using 4-category smoking measures. Here the information criterion was lowest for the five-class model. These latent classes are visualised using stacked bar graphs in Supplementary Figure S3. These five classes are described as (1) early-onset smoking, (2) late-onset smoking, (3) occasional smoking, (4) short-term smoking, and (5) non- smoking. Non- smoking was again the largest class (62.0%). Occasional smoking was the second largest class (13.7%). There were slightly more participants assigned to late-onset smoking (10.8%) compared to early-onset smoking (8.1%). Short-term smoking was the smallest class (5.4%). This shows that the five-class solution using 3-category smoking shows a slightly greater proportion of late and early-onset smoking, and slightly smaller proportion of occasional smoking, compared to the six-class solution using 4-category smoking.

**Latent class growth analysis**

Latent class growth analysis (LCGA) was also used to model the relationship between repeated measures of smoking as this would offer reproducible classes for future studies. This fits an ordered logit model that does assume proportional odds. This differs from LLCA as growth mixture models describe the change in the repeated measures as a function of time for each class but allows for variance in the intercept and shape of this function for individuals within each class. This means far fewer parameters are required for LCGA compared to LLCA, making it more parsimonious, and unequal spacing of measurement waves can be accounted for. LCGA is a constrained form of a growth mixture model as within class variances are set to zero and thus within each class individuals are assumed to have the same predicted trajectory which is determined by class membership.

Supplementary Table S9 show fit statistics for the LCGA models using linear, quadratic, cubic and quartic growth functions to describe change in smoking over time. In all cases the cubic solution showed the lowest BIC and were the only solutions to show a smaller BIC compared to LLCA models. Supplementary Table S10 show fit statistics for the cubic LCGA models fitted with 2 to 7 classes. Conversely to the LLCA models this suggests that when using 4-category smoking the seven-class solution cubic LCGA is better than the six-class solution. Similarly, when using the 3-category smoking measures the 6 or 7 class cubic LCGA model is better than the five-class solution. With this said, in these solutions the smallest classes make up less than 2% of the participants and as such these solutions would not be useable in downstream analyses as this would cause power issues.

Overall, it appears that the most parsimonious LCGA solution with the lowest BIC, and with a smallest class of greater than 4%, is the five-class cubic LCGA model using 3-category smoking variables. The five classes are described as (1) early-onset smoking, (2) late-onset smoking, (3) occasional smoking, (4) short-term smoking, and (5) non- smoking. Non- smoking was again the largest class (61.1%) and occasional smoking the second largest class (17.2%). There were slightly more late-onset smoking participants (10.0%) compared to early-onset smoking (7.2%). Short-term smoking was the smallest class (4.4%). Compared to the five-class LLCA model, the cubic LCGA solution shows a greater proportion of occasional smoking and fewer early-onset and short-term smoking participants. These classes are visualised using bar graphs in Supplementary Figure 3. The shape of the smoking trajectories are very similar to that of the five-class LLCA model.

**Final model chosen for analyses with risk factors of interest**

LLCA models were preferred over LCGA so patterns of change in smoking behaviours are defined by the variable categories, rather than estimated intercepts and slopes. There was minimal reduction of the information criterion from the five- to six-class LLCA models and entropy was greater for the five-class solution, therefore the five-class solution was used in downstream analyses.

### 4. Association analyses

The associations between latent class membership and risk factor measures was assessed using a three-step approach. In step 1, latent classes are estimated using an unconditional LLCA then used to derive class-assignment probabilities. In step 2 participants are assigned to the class for which their probability is greatest creating a non-latent classification. In step 3, measurement error inherent in this classification is quantified and used to reproduce latent classes using a set of logit constraints. This method maintains some uncertainty in the “most likely” class assignments. In Step 3 latent cell counts were derived and used to carry out univariable analyses and calculate odds ratios (OR) and confidence intervals (CI) used to determine the strength and direction of the associations between risk factors and latent classes of smoking. P values were not used to determine significance given sample size varied by the risk factor measure and comparison in question given each latent class had a different number of participants and not all participants had available data for every measure. Only risk factors with an appropriate sample size (Bujang et al., 2018) were investigated (Supplementary Figure S1). To orient these comparisons each pair of classes were ordered in terms of what seemed the more harmful behaviour, namely earlier onset or more frequent or sustained use.

Maternal cotinine was not associated with all smoking latent classes compared to non-smoking. Parental smoking was not associated with all sustained smoking classes compared to short-term smoking. Other parental substance use was not associated with all latent classes of regular compared to occasional smoking, nor sustained compared to short-term smoking. Parental mental health, education, socioeconomic position (SEP), and neighbourhood deprivation were not associated with all smoking latent classes compared to non-smoking. Parental education was not associated with all classes of sustained smoking compared to short-term smoking. The participant’s sex and neighbourhood deprivation were not associated with all smoking latent classes compared to non-smoking. Risk factors related to the participant’s employment were not associated with all classes of sustained compared to short-term smoking. The participant’s mental health was not associated with all latent classes of regular smoking compared to occasional smoking. Adverse childhood experiences (ACEs) or trauma were not associated with all classes of sustained compared to short-term smoking.

**Consistent hits**

85 measures of 31 risk factors were associated with all latent classes of smoking compared to non-smoking. 89 measures of 36 risk factors were associated with all latent classes of regular compared to occasional smoking. 30 measures of 22 risk factors were associated with all latent classes of sustained compared to short-term smoking. 161 measures of 54 risk factors were associated with early-onset compared to late-onset smoking. These are discussed below.

**Family and peer substance use**

Generally maternal smoking increased odds of any smoking, regular smoking, and early-onset smoking compared to non-smoking, occasional smoking and late-onset smoking respectively. However, participants whose mother started smoking in adulthood showed *decreased* odds of early-onset compared to late-onset smoking (18y), and occasional smoking compared to non-smoking (18, 22y). Participants whose mother had stopped smoking (8, 11y) showed decreased odds of regular compared to occasional smoking, and early- compared to late-onset smoking. Participants whose mother had used cannabis showed increased odds of any smoking compared to non-smoking, and early- compared to late-onset smoking. Participants whose mother drank alcohol more than once per week in infancy (1y) showed increased odds of any smoking compared to non-smoking, while later measures (5, 7y) decreased odds of early- compared to late-onset smoking.

Participants whose mother’s partner reported smoking, or who grew up in a household with smokers, showed increased odds of regular compared to occasional smoking, and early- compared to late-onset smoking. Compared to non-smoking, paternal smoking (2, 4, 9, 11y), and household smoking (2, 3y) increased odds of regular smoking, but decreased odds of occasional smoking.

Peer smoking and peer cannabis use increased odds of more harmful smoking patterns across all comparisons. Peer alcohol consumption in adolescence increased odds of any smoking compared to non-smoking (13, 14, 16y), early- compared to late-onset smoking (10, 13, 14, 16y) and early-onset compared to short-term smoking, but decreased odds of late-onset smoking compared to short-term smoking (16y). Having friends who get drunk (20y) or have an alcohol problem (20y) increased odds of any smoking compared to non-smoking while the former decreased odds of regular compared to occasional smoking. Having friends who had been offered drugs in adolescence or later used drugs increased odds of any smoking compared to non-smoking, early- compared to late-onset smoking and early-onset compared to short-term smoking, but decreased odds of late-onset smoking compared to short-term smoking.

**Family sociodemographic factors**

Participants whose mother reported a lower household income showed increased odds of regular compared to occasional smoking (<£2,100 per month, 18y), and early- compared to late-onset smoking (<£360 per week, 11y). Participants whose mother did not own their home or were not married showed increased odds of regular compared to occasional smoking, sustained compared to short-term smoking, and early- compared to late-onset smoking.

Participants whose parents were not economically active (employed or in education/training), mother had fewer qualifications (<O-level), or mother’s partner worked in routine occupations (Registrar General's Social Classes; RGSC) at birth showed increased odds of early- compared to late-onset smoking. GCE Ordinary Levels (AKA O Levels) were exams taken by UK students typically around the age of 16 after completing their compulsory education. This was replaced by GCSEs (General Certificate of Secondary Education) in the late 1980’s. However, participants whose mother was not economically active in adulthood (18y) showed *decreased* odds of sustained compared to short-term smoking. Participants whose mother’s partner had fewer qualifications (0y) showed increased odds of regular compared to occasional smoking.

Participants whose mother lived in more deprived areas with higher (top 2 quintiles) Index of multiple deprivation (0, 1, 2y) and Townsend deprivation scores (1, 2, 7y) showed increased odds of regular compared to occasional smoking and sustained compared to short-term smoking (10y). Participants whose mother lived in more rural areas (12y) showed decreased odds of early- compared to late-onset smoking.

**Individual lifestyle factors**

Participants with a higher BMI (>median) showed increased odds of regular smoking compared to non-smoking (14y) and regular compared to occasional smoking (9, 10, 13, 14, 16, 24y) but decreased odds of occasional smoking compared to non-smoking (14y), sustained compared to short-term smoking (16y) and early- compared to late-onset smoking (18y). Participants who exercised less than weekly in late adolescence showed increased odds of any smoking compared to non-smoking (18y) and early- compared to late-onset smoking (17, 18y) while a later measure (22y) decreased odds. Participants with fewer days (<5) of moderate-to-vigorous exercise (MVPA) showed increased odds of any smoking compared to non-smoking (11, 14y), regular compared to occasional smoking (16y), and early- compared to late-onset smoking (14y). Participants who reported not being part of a club or playing sports in childhood (11y) showed decreased odds of sustained compared to short-term smoking. In adolescence (14y) this decreased odds of late-onset smoking, but increased odds of early-onset smoking, compared to short-term smoking and early- compared to late-onset smoking. Participants who slept for fewer hours (<10) in childhood (11y) showed decreased odds of early- compared to late-onset smoking while in adolescence (16y) fewer hours (<8), increased odds. Fewer hours (<8) asleep in adulthood (25y) increased odds of sustained compared to short-term smoking.

Participants who drank alcohol more than weekly showed increased odds of any smoking compared to non-smoking (13, 14, 16, 17, 18, 20, 22, 24y) and regular compared to occasional smoking (16, 18y). In adolescence alcohol consumption increased odds of early- compared to late-onset smoking (13, 14, 16, 17, 18y) while later measures in adulthood decreased odds (20, 22, 24, 28y). Participants who reported using cannabis showed increased odds of any smoking compared to non-smoking (14, 16, 17, 18, 20, 22, 24, 28y), regular compared to occasional smoking (16, 18y), and early- compared to late-onset smoking (14, 16, 17, 18, 20, 24y). However, cannabis use (16y) or being offered drugs (14y) increased odds of early-onset, but decreased odds of late-onset smoking, compared to short-term smoking. Later cannabis use (28y) increased odds of sustained compared to short-term smoking. Participants who had been offered drugs (13, 14y) showed increased odds of any smoking compared to non-smoking and early- compared to late-onset smoking. Participants who reported ever using drugs showed increased odds of more harmful smoking patterns across all comparisons.

**Individual sociodemographic factors**

Participants assigned female at birth showed decreased odds of sustained compared to short-term smoking and late-onset compared to occasional smoking, but increased odds of early-onset and short-term smoking compared to occasional smoking, and early- compared to late-onset smoking.

Participants who did not plan to stay in education showed increased odds of any smoking compared to non-smoking (16y) and early- compared to late-onset smoking (14, 16, 18y). Participants with lower educational qualifications (18, 20, 26y), fewer years of education (26y) or were not in education (22y) showed increased odds of early- compared to late-onset smoking. Participants who were currently studying at below A-levels (18y) showed decreased odds of early- compared to late-onset smoking. A-levels, or "Advanced Level" qualifications, typically follow the completion of GCSE or equivalent qualifications, usually by students in the age range of 16 to 18. Participants with lower qualifications showed increased odds of regular smoking compared to non-smoking (20y), regular compared to occasional smoking (18, 20y) and early-onset compared to short-term smoking (18, 20y), but decreased odds of late-onset smoking compared to short-term smoking (18, 20y), and occasional smoking compared to non-smoking (20y). Participants who were not in education or training showed increased odds of regular smoking compared to non-smoking (21y), regular compared to occasional smoking (20, 21y) and early- compared to late-onset smoking (18, 20y), but decreased odds of occasional smoking compared to non-smoking (21y). Participants who were not economically active or worked in supervisory, technical or routine occupations (National Statistics Socio-Economic Classification; NS-SEC) showed increased odds of regular compared to occasional smoking (23y). Participants who worked nights or shifts showed increased odds of early- compared to late-onset smoking (25y). Participants who earned a higher income (>£1500 per month) in adulthood (25y) showed decreased odds of regular smoking compared to non-smoking and regular compared to occasional smoking, but increased odds of occasional smoking compared to non-smoking.

Participants who lived in areas with higher Index of Multiple Deprivation (IMD) or Townsend deprivation scores (ReStore National Centre for Research Methods; Townsend et al, 1988; Morgan and Baker, 2006) showed increased odds of sustained compared to short-term smoking (11, 12y) and early- compared to late-onset smoking (13, 14, 18y). Participants who lived in more rural areas (10, 13, 14y) showed decreased odds of early- compared to late-onset smoking.

**Mental health and other factors**

Participants with mothers who had a mental health condition showed increased odds regular compared to occasional smoking (5y) and early- compared to late-onset smoking (0y), but decreased odds of sustained compared to short-term smoking (5y). Participant’s whose mother reported feeling depressed (2y) showed increased odds of regular compared to occasional smoking. Participants whose mother reported having more bad than good days (8w), or whose maternal grandmother had a mental health condition (0y), showed increased odds of early- compared to late-onset smoking. Participants whose mother’s partner had a mental health condition (6y) showed decreased odds of sustained compared to short-term smoking.

Participants who stated feeling miserable in the Development and Well-Being Assessment (DAWBA, 16y) (Goodman et al., 2000), had a mental health condition (22y), took mental health medication (28y), had higher Clinical Interview Schedule-Revised (CIS-R) scores (>12, 18y) (Lewis et al., 1992), symptoms of generalised anxiety (24y) or moderate depression (24y) showed increased odds of any smoking compared to non-smoking. Participants with higher CIS-R scores (18y), symptoms of chronic fatigue (18y), generalised anxiety (18y), mild (18y) or moderate (18y) depression, higher Mood and Feelings Questionnaire (MFQ) scores (>12, 14y) (Angold et al., 1995), or stated losing interest in thing they normally enjoy (16y) showed increased odds of early- compared to late-onset smoking. Participants with lower Warwick-Edinburgh Mental Wellbeing Scale (WEMWS) scores (<42) (Tennant et al., 2007) showed increased odds of sustained compared to short-term smoking (18y) and early- compared to late-onset smoking (23y).

Participants who had a baby (24y), had been pregnant (21, 22, 24y), or were a parent (22, 23, 25, 28y) showed increased odds of early- compared to late-onset smoking. Participants who had been pregnant (21y) showed increased odds of any smoking compared to non-smoking. Participants who were parents showed increased odds of regular compared to occasional smoking (22, 23, 24, 28y) and early-onset compared to short-term smoking (22y), but decreased odds of late-onset smoking compared to short-term smoking (22y).

Participants who had experienced more (>2) adverse childhood experiences ([ACEs], 0-16y) showed increased odds of any smoking compared to non-smoking and regular compared to occasional smoking. Participants who had experienced trauma showed increased odds of any smoking compared to non-smoking (0-17, 11-17y), regular compared to occasional smoking (0-17y), and early- compared to late-onset smoking (0-5, 0-17, 11-17y).

### 5. R Packages

R packages used include xlsx (Dragulescu and Arendt, 2020), dplyr (Wickham et al., 2022a), haven (Wickman et al., 2022b), ggplot2 (Wickman, 2016), ggrepel (Slowikowski, 2022), ggpubr (Kassambara, 2023), plotly (Sievert, 2020), reshape2 (Wickham, 2007), labelled (Larmarange, 2022), glue (Hester an Bryan, 2022), circlize (Gu, 2014), ComplexHeatmap (Gu, 2022) and htmlwidgets (Vaidyanathan et al., 2023).
